# Visuomotor paired associative stimulation enhances corticospinal excitability in post-stroke patients with upper-limb hemiparesis: A proof-of-principle study

**DOI:** 10.1101/2024.08.27.24312576

**Authors:** Michela Picardi, Giacomo Guidali, Antonio Caronni, Viviana Rota, Massimo Corbo, Nadia Bolognini

**Author notes:** ***Corresponding authors:*** Nadia Bolognini, PhD Department of Psychology, University of Milano-Bicocca, Piazza dell’Ateneo Nuovo 1, Milan, Italy Giacomo Guidali, PhD Department of Psychology, University of Milano-Bicocca, Piazza dell’Ateneo Nuovo 1, Milan, Italy. ***These authors contributed equally to this work*** (Michela Picardi, Giacomo Guidali).

## Abstract

We assess the effectiveness of a visuomotor *paired associative stimulation* (vm-PAS) protocol targeting the Action Observation Network (AON) in chronic post-stroke patients with upper-limb mild hemiparesis. Vm- PAS consisted of hand-grasping action observation stimuli repeatedly paired with transcranial magnetic stimulation (TMS) pulses over the ipsilesional primary motor cortex (M1).

Fifteen post-stroke patients underwent a session of the vm-PAS and, as a control, of the standard excitatory PAS (M1-PAS), during which slow-rate electrical stimulation of the paretic limb was paired with M1-TMS. Before and after each PAS, we assessed corticospinal excitability (CSE), short-interval intracortical inhibition (SICI), and paretic wrist’s voluntary movements.

The two protocols induce distinct muscle-specific CSE enhancements: vm-PAS increases motor-evoked potentials (MEPs) recorded from the paretic *first dorsal interosseous* muscle. Conversely, M1-PAS increases MEPs recorded from the electrically stimulated *extensor carpi radialis* muscle. Vm-PAS efficacy correlates with hemiparesis chronicity: the higher the time elapsed since the stroke, the greater vm-PAS effects on CSE. Neither protocol affected SICI or wrist movements.

Our results suggest that vm-PAS leads to muscle-specific enhancements of CSE in post-stroke patients, highlighting its potential for driving post-stroke motor recovery. These findings show the efficacy of a cross-modal PAS protocol targeting the AON in an injured motor system.

## 1. INTRODUCTION

Motor impairments after a stroke are the primary cause of disability in adults (Mayo et al., 2002). Six months after a stroke, between 50% and 70% of patients still suffer from upper extremity disabilities, such as paresis and spasticity, for which the chances of recovery are drastically limited even despite continuing intensive rehabilitation treatments (Lang et al., 2016; Pollock et al., 2014). Non-invasive brain stimulation may help maximize post-stroke motor recovery by modulating cortical excitability and brain plasticity even in the chronic phase of illness (Ahmed et al., 2011; Anwer et al., 2022; Di Pino et al., 2014; Su & Xu, 2020).

Among these techniques, paired associative stimulation (PAS) has attracted neuroscientific and clinical research because it can target specific cortico-peripheral and cortico-cortical pathways in a timing-dependent way (Guidali et al., 2021b, 2021a). PAS repeatedly pairs time-locked peripheral and cortical stimulations, the last represented by a transcranial magnetic stimulation (TMS) pulse over the cortical site of interest. This repeated contingency promotes the functional reorganization of brain networks through mechanisms of Hebbian associative plasticity within the target area/circuit (Suppa et al., 2017). The classic version of the PAS (here, M1-PAS); combines TMS pulses applied to the primary motor cortex (M1) with peripheral electrical stimulation (Stefan et al., 2000; Wolters et al., 2003). Its effectiveness in inducing long-term potentiation or depression of corticospinal excitability (CSE) is well-documented in healthy subjects but, to a lesser extent, in the clinical population (Baroni et al., 2024; Wischnewski & Schutter, 2016). In the case of stroke patients, the presence of hemiparesis or hemianesthesia may compromise the effectiveness of the peripheral electrical stimulation required to activate the motor system through the afferent pathway, in turn impacting the induction of associative plasticity (Ferris et al., 2018; Silverstein et al., 2019). To overcome this issue, accessing the motor system through other afferent pathways, such as via the visual system, may be useful, considering that they might be preserved in patients with hemiparesis. This indirect access to the motor system could be achieved by taking advantage of a novel, visuomotor version of the PAS that pairs action observation stimuli with TMS pulses over M1 (the so-called mirror-PAS - Guidali et al., 2020). Indeed, in healthy individuals, this repeated coupling was shown to promote associative plasticity mechanisms within M1, likely driven by the recruitment of the Action Observation Network (AON - Kemmerer, 2021; Rizzolatti & Craighero, 2004), with benefits detectable at the neurophysiological and behavioral level (Guidali et al., 2020, 2025; Guidali, Picardi, Gramegna, et al., 2023; Guidali & Bolognini, 2025). However, to date, the potential of visuomotor versions of the PAS for post-stroke motor deficits has not yet been determined, although substantial clinical improvements can be achieved through AON activation, as demonstrated by rehabilitation approaches such as the mirror box illusion and action observation therapies (Thieme et al., 2018; Zhang et al., 2023).

In this scenario, the present proof-of-principle study explored the effectiveness of a visuomotor PAS (vm-PAS) applied over the ipsilesional motor cortex during the passive observation of hand grasping actions in a sample of post-stroke patients with chronic upper limb hemiparesis, collating its effect to those induced by an excitatory version of the conventional M1-PAS (Baroni et al., 2024). This latter protocol, pairing electric stimulations of the paretic limb with TMS pulses over the injured M1, has already been proven effective in the stroke population (Castel-Lacanal et al., 2007, 2009) and should act as a control condition for the vm-PAS, allowing us to compare the magnitude of the modulations induced by the two protocols within the same cohort of hemiparetic patients.

Patients underwent a single session of both PAS protocols, assessing their efficacy at the neurophysiological level (primary outcome: corticospinal excitability - CSE - and short-interval intracortical inhibition – SICI) and behavioral level (secondary outcome: voluntary wrist movement of the paretic limb). CSE and SICI were measured by recording motor-evoked potentials (MEPs) from three different muscles of the paretic upper limb (i.e., *extensor carpi radialis* – ECR, *flexor carpi radialis* – FCR, and *first dorsal interosseus* – FDI) to verify the extent to which the vm-PAS aftereffects are selective for the muscles involved in the observed action, as predicted by the functionality of the AON (e.g., Fadiga et al., 1995; Guidali et al., 2020; Guidali, Picardi, Franca, et al., 2023; Naish et al., 2014).

## 2. MATERIALS AND METHODS

### 2.1. Participants

Twenty adults with chronic post-stroke upper-limb hemiparesis were recruited at the Department of Neurorehabilitation Sciences of the Istituto Auxologico Italiano (Capitanio Hospital, Milan, IT), according to the following inclusion criteria: aged between 18-85 years; affected by ischemic or hemorrhagic stroke, cortical or subcortical, in supratentorial site; stroke occurred from at least 4 months; the presence of clinically documented upper limb hemiparesis in the dominant (before stroke) hand; no history of psychiatric, neurological, neuropsychological and musculoskeletal conditions that compromise upper limb function or prevent understanding experimental instructions; no contraindications to TMS (Rossi et al., 2021). All our subjects were outpatients, without interventions occurring parallelly during testing; they all completed a traditional rehabilitation program during the acute stage of the stroke. Two out of 20 patients dropped out for personal issues, and for 3 patients, reliable MEPs could not be evoked in the target muscles at the baseline assessment (see **Study design**). Thus, the final analyzed sample comprised 15 patients with right-hand hemiparesis, whose demographic and clinical characteristics are summarized in **Table 1**.

**Table 1.**
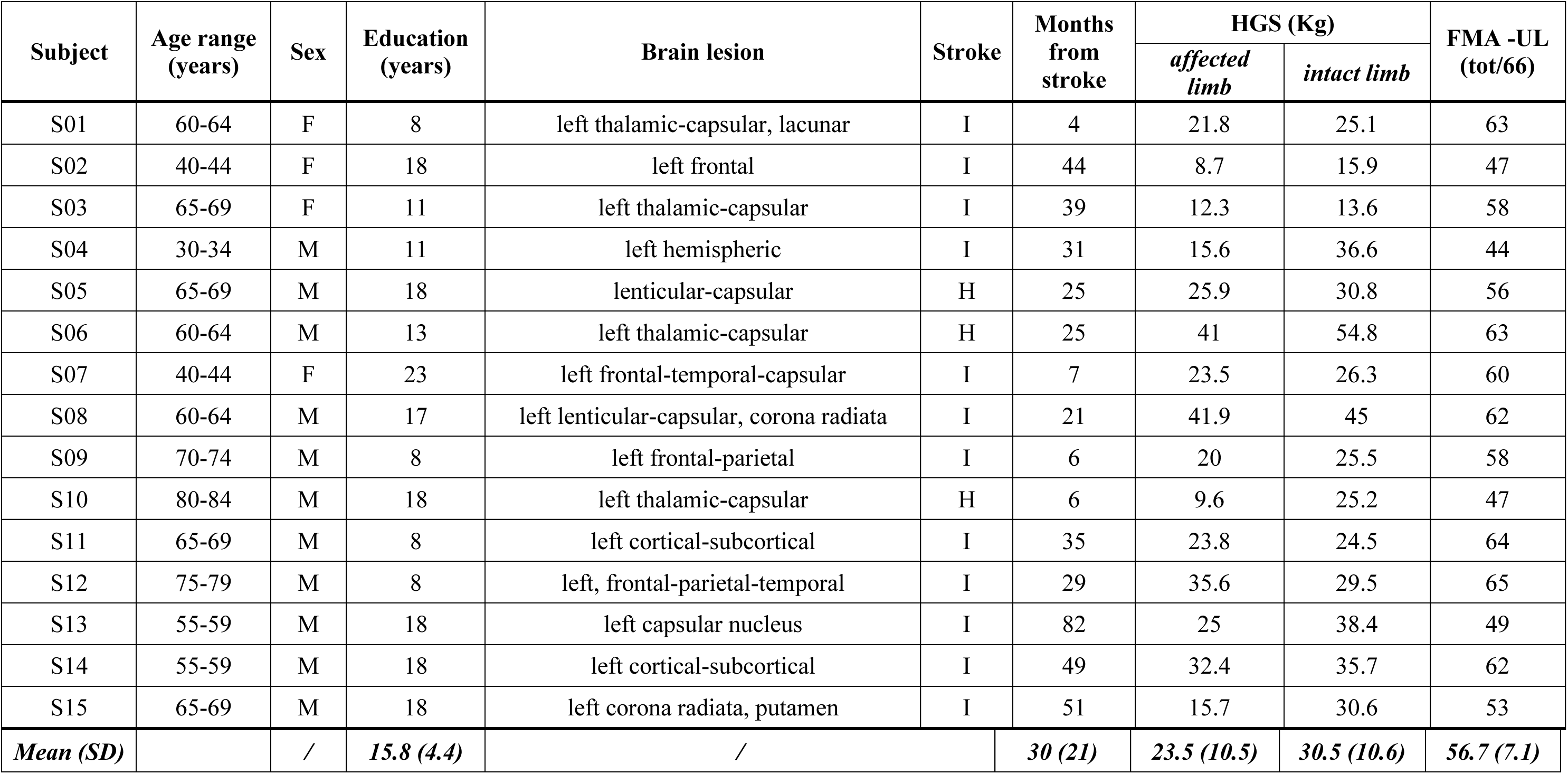
Demographic and clinical characteristics of stroke sample. All patients had a left-sided hemispheric lesion; hence, a right-sided upper limb hemiparesis. Legend: F=female, M=male, I=ischemic stroke, H=hemorrhagic stroke, HGS=Hand Grip Strength, FMA-UL=Fugl-Meyer Assessment Upper Limb, OCS=Oxford Cognitive Scale, SD=standard deviation.

| Subject | Age range (years) | Sex | Education (years) | Brain lesion | Stroke | Months from stroke | HGS (Kg) |  | FMA -UL (tot/66) |
| --- | --- | --- | --- | --- | --- | --- | --- | --- | --- |
|  |  |  |  |  |  |  | *affected limb* | *intact limb* |  |
| S01 | 60-64 | F | 8 | left thalamic-capsular, lacunar | I | 4 | 21.8 | 25.1 | 63 |
| S02 | 40-44 | F | 18 | left frontal | I | 44 | 8.7 | 15.9 | 47 |
| S03 | 65-69 | F | 11 | left thalamic-capsular | I | 39 | 12.3 | 13.6 | 58 |
| S04 | 30-34 | M | 11 | left hemispheric | I | 31 | 15.6 | 36.6 | 44 |
| S05 | 65-69 | M | 18 | lenticular-capsular | H | 25 | 25.9 | 30.8 | 56 |
| S06 | 60-64 | M | 13 | left thalamic-capsular | H | 25 | 41 | 54.8 | 63 |
| S07 | 40-44 | F | 23 | left frontal-temporal-capsular | I | 7 | 23.5 | 26.3 | 60 |
| S08 | 60-64 | M | 17 | left lenticular-capsular, corona radiata | I | 21 | 41.9 | 45 | 62 |
| S09 | 70-74 | M | 8 | left frontal-parietal | I | 6 | 20 | 25.5 | 58 |
| S10 | 80-84 | M | 18 | left thalamic-capsular | H | 6 | 9.6 | 25.2 | 47 |
| S11 | 65-69 | M | 8 | left cortical-subcortical | I | 35 | 23.8 | 24.5 | 64 |
| S12 | 75-79 | M | 8 | left, frontal-parietal-temporal | I | 29 | 35.6 | 29.5 | 65 |
| S13 | 55-59 | M | 18 | left capsular nucleus | I | 82 | 25 | 38.4 | 49 |
| S14 | 55-59 | M | 18 | left cortical-subcortical | I | 49 | 32.4 | 35.7 | 62 |
| S15 | 65-69 | M | 18 | left corona radiata, putamen | I | 51 | 15.7 | 30.6 | 53 |
| **Mean (SD)** |  | / | **15.8 (4.4)** | / |  | **30 (21)** | **23.5 (10.5)** | **30.5 (10.6)** | **56.7 (7.1)** |

We determined the sample size through an a-priori within-subjects repeated measures ANOVA (rmANOVA) using the software G*Power 3.1 (Faul et al., 2009) and selecting as the target effect size value the smallest one among those found in our previous studies using visuomotor PAS protocols (Guidali et al., 2020, 2025; Guidali, Picardi, Gramegna, et al., 2023), which corresponded to a partial eta squared (*η_p2_*) of .16 [Experiment 1 in Guidali, Picardi, Gramegna, et al. (2023)]. The power analysis (alpha: *p* = .05; statistical power = .8, actual power = .85) showed a recommended sample size of at least 14 participants to achieve enough statistical power for detecting PAS effects.

All patients provided their written informed consent to participate in this study, which was performed following the standards of the Declaration of Helsinki (2013) and obtained approval by the local Ethics Committee (protocol n° 25C212) of the Istituto Auxologico Italiano. The study was registered on ClinicalTrial.gov in date 13/03/2023 (n° NCT05766059; https://clinicaltrials.gov/study/NCT05766059).

### 2.2. TMS and electromyographic (EMG) recording

TMS pulses were delivered with a figure-of-eight coil (Ø= 70 mm) connected to a monophasic stimulator (Magstim 200^2^, The Magstim Company Ltd, Whitland, UK) over the ECR hotspot of the ipsilesional (left in every patient) M1. The hotspot was defined as the stimulation site consistently inducing the largest MEPs in the paretic ECR muscle, with visually detectable wrist extension at intensities above the resting motor threshold (rMT). Importantly, in all our patients, the supra-threshold stimulation of this point (see **Neurophysiological assessment**) also elicited MEPs in the other two muscles of interest (FDI and FCR). The individual rMT was determined in each PAS session using the parameter estimation by sequential testing procedure, a maximum-likelihood threshold-hunting procedure optimized for rMT detection (Awiszus, 2003; Dissanayaka et al., 2018). The mean rMT was 55.8 ± 18.8% in the vm-PAS session and 55.5 ± 19.2% in the M1-PAS session (*t*_14_ = .51, *p* = .62). ECR representation in M1 was targeted by positioning the coil 45° to the midline, tangential to the scalp, inducing a posterior-anterior current flow. Neuronavigation was performed to monitor stable coil positioning during the TMS stimulation (SofTaxic Optic 3.4, EMS, Bologna, IT).

EMG was recorded from paretic’s ECR, FDI, and FCR muscles with surface electrodes (15 × 20 mm Ag-AgCl pre-gelled surface electrodes, Friendship Medical, Xi’an, China) in a tendon-belly arrangement. The ground electrode was placed over the head of the ulna. The EMG signal was monitored online on a computer screen. Before data acquisition, the experimenter did a visual inspection to guarantee that the background noise from the three muscles was < 30 µV. The raw signal was sampled (5000 Hz), amplified, notch filtered (bandpass 10-1000 Hz), and stored for offline analysis by using Signal software (6.0 version, Cambridge Electronic Devices, Cambridge, UK) connected to a 1902 amplifier and CED Power1401 A/D converter.

### 2.2. PAS protocols

During both PAS protocols, TMS pulses were applied over the ECR representation in the M1 of the injured left hemisphere.

In the vm-PAS (**Figure 1a**), each TMS pulse (intensity = 120% rMT) was paired with a visual stimulus of movement created by presenting two frames: a first (‘static’) frame lasting 4250 ms, depicting a hand viewed from an egocentric perspective and a bottle, which was followed, after an interstimulus interval (ISI) of 25 ms, by a second (‘action’) frame of 750 ms showing the same hand grasping the bottle. The rapid presentation of the second frame gave the perception of the hand’s apparent motion, performing a reaching-to-grasp action. This type of visual stimulus is suitable for the clinical population, being typically exploited in action observation therapies for stroke patients (Franceschini et al., 2012; Tropea et al., 2023). One hundred eighty paired visual and TMS stimuli were delivered at .2 Hz (Guidali et al., 2020, 2025; Guidali, Picardi, Gramegna, et al., 2023; Guidali & Bolognini, 2025). To ensure that patients paid attention to the visual stimuli, we asked them to count how many times the bottle was grasped by the depicted hand. E-Prime software (3.0, Psychology Software Tool, Inc., Pittsburgh, USA) controlled the timing of the stimuli and TMS administration.

**Figure 1.**
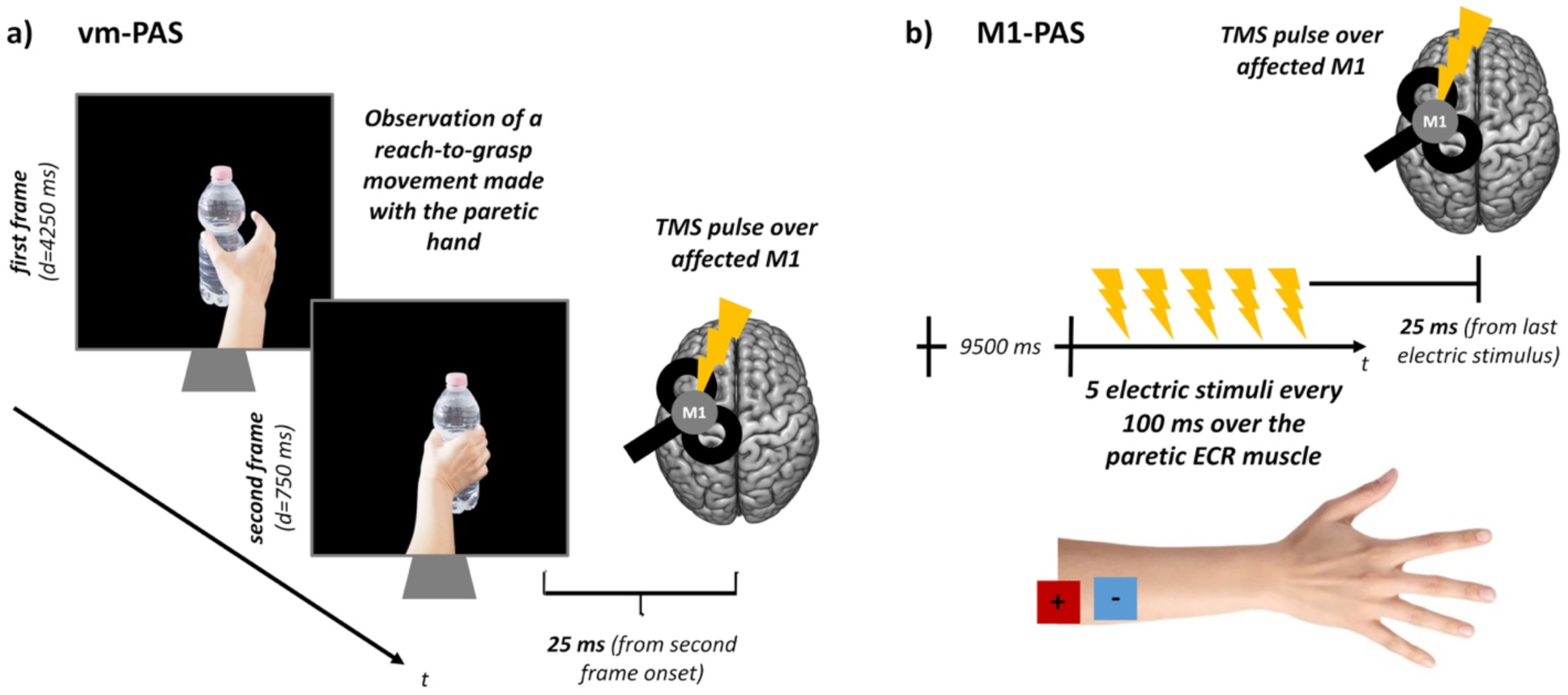
PAS protocols. Schematic illustration of the vm-PAS (**a**) and the M1-PAS (**b**) protocols administered in the present work.

The M1-PAS (**Figure 1b**) consisted of trains of ECR electrical stimulation (5 stimuli at 10 Hz with square waves of 1 ms; intensity adjusted to produce the minimal visible ECR contraction) paired with the TMS over M1 at 120% rMT. The peripheral stimulation of the ECR was chosen based on previous M1-PAS literature on stroke patients (Castel-Lacanal et al., 2007, 2009; Tarri et al., 2018) and given the importance of this muscle in hand motor recovery since it stabilizes the wrist and ensures an effective hand grip (Kamper et al., 2003). TMS pulses were applied with an ISI of 25 ms from the last electric stimuli of the train. One hundred eighty paired stimulations were administered at .1 Hz (Carson & Rankin, 2018; Castel-Lacanal et al., 2007, 2009). Electrical stimulation was delivered with a constant current stimulator (Digitimer DS7AH, Digitimer Ltd., Welwyn, UK) and a belly-belly electrode arrangement. During the M1-PAS, patients were asked to count the number of peripheral electrical stimuli received (Stefan et al., 2004). Electric stimuli and TMS were delivered under computer control (Signal software).

### 2.3. Neurophysiological assessment

During the neurophysiological assessment, patients were seated in a comfortable position with both upper limbs at rest on the table (the paretic upper limb was in a pronated position and supported on a cushion); they were instructed to keep eyes fixation on a red asterisk appearing on the PC screen in front of them.

CSE was assessed by administering 30 TMS pulses at 120% rMT over the ECR representation in the ipsilesional M1, with an inter-trial interval between 4000 and 5000 ms. SICI was assessed using a dual-coil device (Magstim BiStim^2^, The Magstim Company Ltd, Whitland, UK) by applying a conditioning TMS stimulus at 80% rMT over the ECR representation in ipsilesional M1 followed, after 2 ms, by a test pulse at 120% rMT over the same cortical site (Kujirai et al., 1993). Fifteen paired and 15 single pulses were randomly delivered every 10 sec to record conditioned and unconditioned MEPs.

### 2.4. Measurement of voluntary wrist movement

The wrist’s voluntary movement of the paretic limb was captured by using an inertial movement unit (IMU) (Martino Cinnera et al., 2024), consisting of an integrated unit composed of a 3-axis gyroscope, a 3-axis accelerometer, and a magnetometer (G-sensor, BTS Bioengineering Corp., Quincy, USA). Patients sat comfortably with their affected forearm stabilized on the chair’s armrest. The wrist was free to move, with the forearm pronated for the extension movement and supinated for the flexion movement assessment. The IMU was fixed to the dorsal surface of the hand at the level of the third metacarpal bone and tightly secured to the participant’s skin with adhesive tape. After a familiarization period, the subject was asked to perform two ballistic movements as fast as possible: extension and flexion of the wrist. Each movement was performed 5 times, with 5 sec of rest between them. The wrist movements were recorded using the BTS software (G-studio).

### 2.5. Study design

Before taking part in the experiment, on a separate day from PAS sessions, patients underwent a baseline assessment comprising the Fugl-Meyer Assessment Upper Limb scale – FMA-UL (Fugl-Meyer et al., 1975), the Hand Grip Strength test – HGS (Hamilton et al., 1992), and the Oxford Cognitive Scale – OCS (Mancuso et al., 2016) (see **Table 1** and **Supplemental Table 1**). Experimenters also checked if MEPs greater than 100 µV can be reliably recorded at supra-threshold intensities in the paretic limb from the three target muscles.

The experiment consisted of two counterbalanced within-subject sessions during which patients underwent the vm-PAS and the M1-PAS, whose order was randomized across patients. Immediately before and after each PAS, patients underwent the neurophysiological assessment (i.e., CSE and SICI) and the measurement of the paretic limb’s voluntary wrist movement. The neurophysiological and behavioral assessment order was kept fixed for all patients.

Each patient underwent the two PAS at the same time of the day (i.e., in the morning or the afternoon) to control for any potential influence of circadian rhythms (Sale et al., 2007), with at least 72 hours between the two sessions. Each PAS session lasted about 1 hour and a half.

### 2.6. Statistical analyses

Data analyses were conducted using the software Jamovi with the package GAMLj [v. 2.4.12; (The Jamovi Project, 2025)] and R Studio (R Core Team, 2019). In every analysis, statistical significance was set with *p* ≤ .05, and significant effects were explored with post-hoc tests with the Bonferroni correction for multiple comparisons. Variables are reported as mean ± standard error (SE). Detailed results from all the analyses can be found in **Supplemental Tables 3-8**. The study’s datasets and scripts are available at Open Science Framework – OSF: https://osf.io/xgn7f/.

#### 2.6.1. Neurophysiological assessment

MEPs were preprocessed with Signal software, exploiting the standard pipeline used in our laboratory (Guidali, Picardi, Gramegna, et al., 2023). Trials with muscular artifacts or background noise deviating from 50 µV in the 100 ms before the TMS pulse were automatically excluded from the analysis. Then, MEP peak-to-peak amplitude was calculated in each trial in the time window between 10 ms and 60 ms from the TMS pulse. For CSE assessment, trials with MEP amplitude smaller than 50 µV were excluded from the analyses.

Visual inspection of the QQ-plot and skewness/kurtosis values showed that raw MEP amplitudes were not normally distributed during CSE assessments. Base-10 log transformation made the distribution closer to normality (see **Supplemental Table 2**); hence, MEPs were transformed accordingly and then analyzed using linear mixed models (LMMs). Each muscle (ECR, FDI, and FCR) was analyzed separately with within-subjects factors ‘PAS’ (vm-PAS, M1-PAS), ‘Time’ (pre-PAS, post-PAS), and their interaction as fixed effects; the intercept as the random one; and the patient as the cluster variable.

For the SICI, we calculated the ratio between conditioned and unconditioned MEPs [where values < 1 indicate inhibition during conditioned trials (Kujirai et al., 1993)], and we analyzed it with a series of ‘PAS’ (vm-PAS, M1-PAS) X ‘Time’ (pre-PAS, post-PAS) rmANOVAs separated for each muscle. Two patients were excluded from SICI analyses due to recording issues during its assessment. To assess whether CSE was still modulated during the SICI, we also conducted an LMM on (log-transformed) MEPs recorded during this assessment (see **Supplemental Analysis 1**).

#### 2.6.2. Voluntary wrist movement

IMU signal extraction was performed using Signal software. For each of the 5 wrist extension and flexion movements, the following measures were manually scored, and their median was considered as the dependent variable: range of motion (ROM, °); maximum peak of angular velocity (AV, °/s); peak of movement’s acceleration (ACC, °/s^2^), calculated as the ratio between the maximum peak of angular velocity and its latency. Then, these parameters were analyzed through a rmANOVA with within-subjects factors ‘PAS’ and ‘Time’, separated for extension and flexion movements.

#### 2.6.3. Correlations between PAS modulation and patients’ clinical profile

We run Spearman’s correlation analyses to explore the influence of (a) stroke chronicity, (b) severity of motor impairment (i.e., scores at FMA-UL and HGS for the affected hand; **Table 1**), (c) patient’s age and educational level, (d) M1 excitability (i.e., rMT), and (e) voluntary wrist movements (i.e., ROM, AV, ACC) at baseline on significant PAS-induced enhancement of CSE (see **Results**). This was expressed as the ratio between MEPs recorded at rest after and before protocols’ administration: 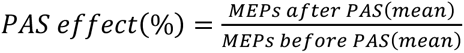. Positive values express MEP enhancement after PAS, while negative ones show MEP depression.

## 3. RESULTS

### 3.1. Neurophysiological assessment

Considering CSE for the ECR muscle, LMM showed a significant main effect of ‘Time’ (*F*_1,1734_ = 21.85, *p* < .001) and ‘PAS’ X ‘Time’ interaction (*F*_1,1734_ = 47.72, *p* < .001): CSE (i.e., log_10_-transformed MEP, µV) significantly increased only after M1-PAS (post-M1-PAS: 2.64 ± .08 *vs.* pre-M1-PAS: 2.53 ± .07, *t*_1734_ = 8.22, *p* < .001; *vs.* pre-vm-PAS: 2.60 ± .06, *t*_1734_ = 2.71, *p* = .041; *vs.* post-vm-PAS: 2.58 ± .08, *t*_1734_ = 4.29, *p* < .001).

Conversely, after the vm-PAS, no modulation of ECR MEPs was found (*t*_1734_ = 1.57, *p* = .7). At the single-subject level, 13 out of 15 patients showed an increase of ECR MEPs amplitude, with an overall mean increase of +33% ± 8.6% (range -9/+104%; **Figure 2a**).

**Figure 2.**
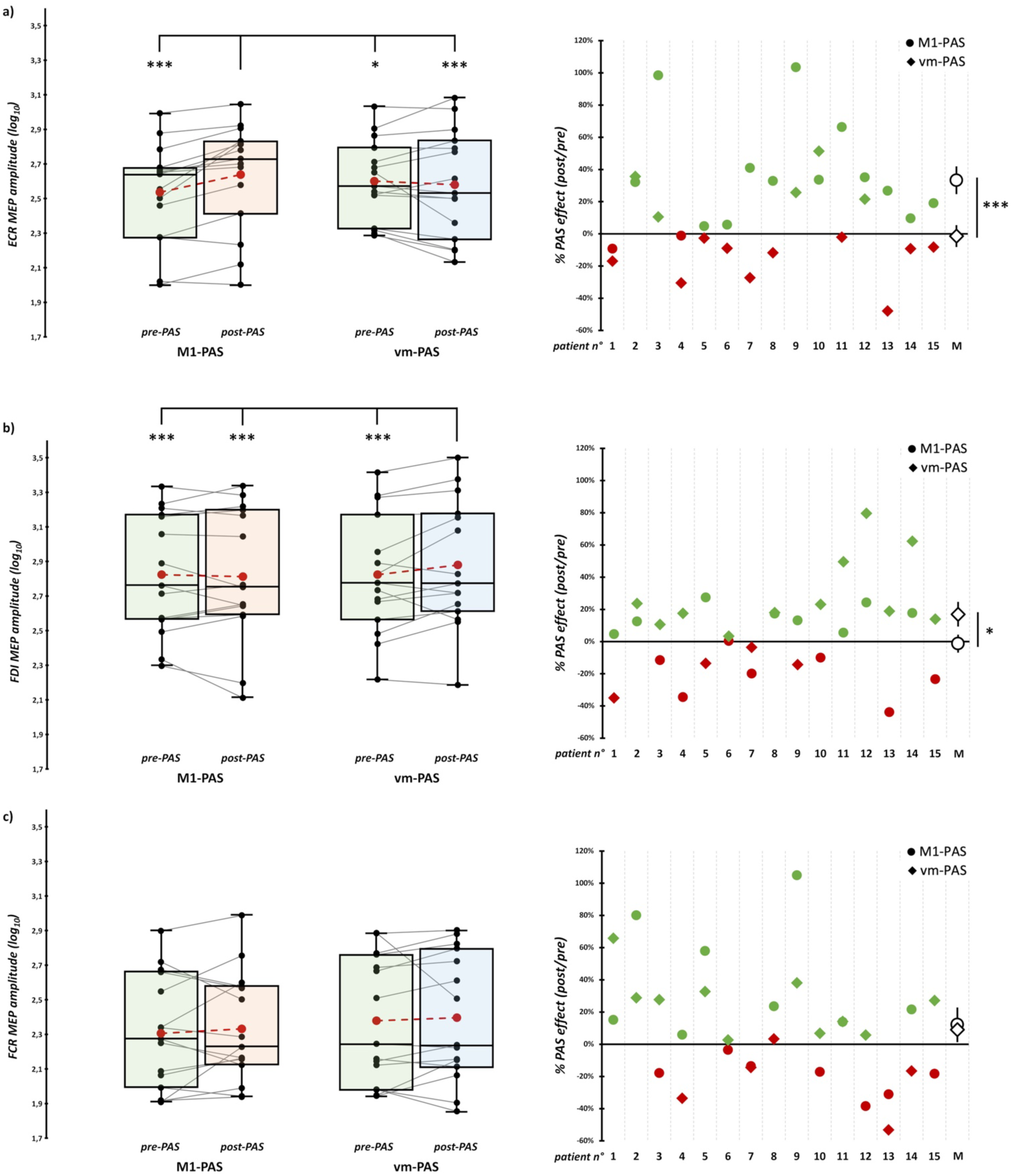
CSE results. Left panels: LMM results from the assessment of CSE for ECR (**a**), FDI (**b**), and FCR (**c**) before (green boxes) and after vm-PAS (blue boxes) and M1-PAS (orange boxes) administration. For ECR, (log_10_- transformed) MEPs significantly increased only after M1-PAS while, for FDI, they increased only after vm-PAS. For FCR muscle, we found that MEPs were greater after both PAS administrations. In the box-and-whiskers plots, red dots and lines indicate the means of the distributions. The center line denotes their median values. Black dots and grey lines depict individual MEP values (N=15). The box contains the 25th to 75th percentiles of the dataset.

For FDI, LMM showed a main effect of ‘PAS’ (*F*_1,1734_ = 14.86, *p* < .001), ‘Time’ (*F*_1,1734_ = 6.57, *p* = .01), and their interaction (*F*_1,1734_ = 15.77, *p* < .001). Post-hoc comparisons showed the opposite pattern than the one found for ECR: CSE increased only after the vm-PAS protocol (post-vm-PAS: 2.88 ± .1 *vs.* pre-vm-PAS: 2.82 ± .09, *t*_1734_ = 4.60, *p* < .001; *vs.* pre-M1-PAS: 2.83 ± .09, *t*_1734_ = 4.53, *p* < .001; *vs.* post-M1-PAS: 2.82 ± .1, *t*_1734_ = 5.55, *p* < .001), while no modulation was found after M1-PAS (*t*_1734_ = 1, *p* = .99). At the single-subject level, 11 out of 15 patients were vm-PAS responders (mean enhancement +17±7.7%; range -35/+80%; **Figure 2b**). Concerning the FCR muscle, we found main effects of factors ‘PAS’ (*F*_1,1734_ = 55.02, *p* < .001 – MEPs recorded during vm-PAS session were greater than in M1-PAS session) and ‘Time’ (*F*_1,1734_ = 5.72, *p* = .017 – MEPs were greater after both PAS administration), but not of their interaction (*F*_1,1734_ = .16, *p* = .688; **Figure 2c**).

Whiskers extend to the largest observation falling within the 1.5 * inter-quartile range from the 1^st^/3^rd^ quartile. Right panels: single-patients MEP enhancement after vm-PAS (rhombi) and M1-PAS (circles) administration. Green rhombi/circles indicate MEP enhancement, while red ones indicate MEP depression. The last row (M) indicates the mean effects of the two PAS. Asterisks indicate significant differences between conditions (* = p < .05, ** = p < .01; *** = p < .001; Bonferroni corrected).

Considering SICI assessment, rmANOVAs showed no statistically significant effects for any of our muscles (all *F*s < .75, all *p*s > .405; **Figure 3; Supplemental Table 6**). This evidence suggests that both protocols have no effect on SICI.

**Figure 3.**
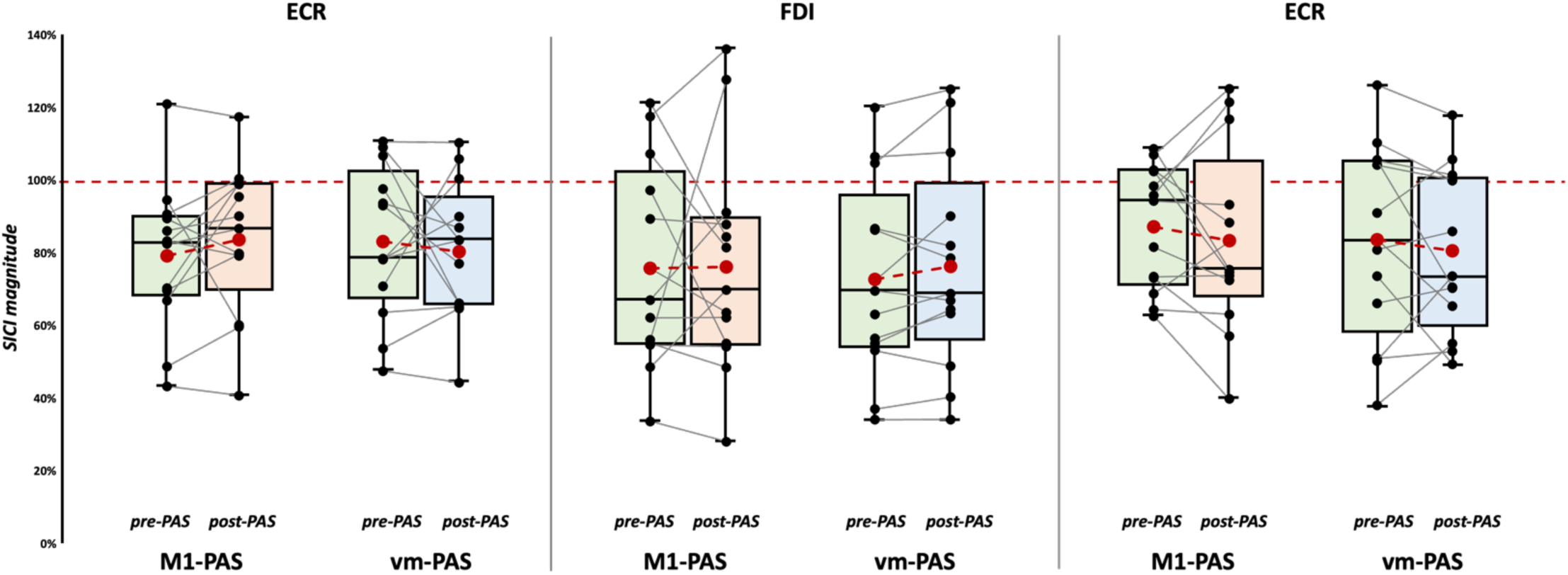
SICI results. Results obtained from the ratio between conditioned and unconditioned MEPs during SICI assessment, before (green boxes) and after vm-PAS (blue boxes) and M1-PAS (orange boxes) administration for ECR (left panel), FDI (middle panel), and FCR (right panel) muscles. No significant effects were found for median ROM, AV, and ACC values. Red dots and lines indicate the means of the distributions. The center line denotes their median values. Black dots and grey lines show individual scores (n=13). The box contains the 25th to 75th percentiles of the dataset. Whiskers extend to the largest observation falling within the 1.5 * inter-quartile range from the first/third quartile.

### 3.2. Voluntary wrist movement

Considering median ROM, AV, and ACC values, every rmANOVA did not show significant main effects or interactions (all *F*s < 3.51, all *p*s > .082; **Figure 4**; **Supplemental Tables 9-10**).

**Figure 4.**
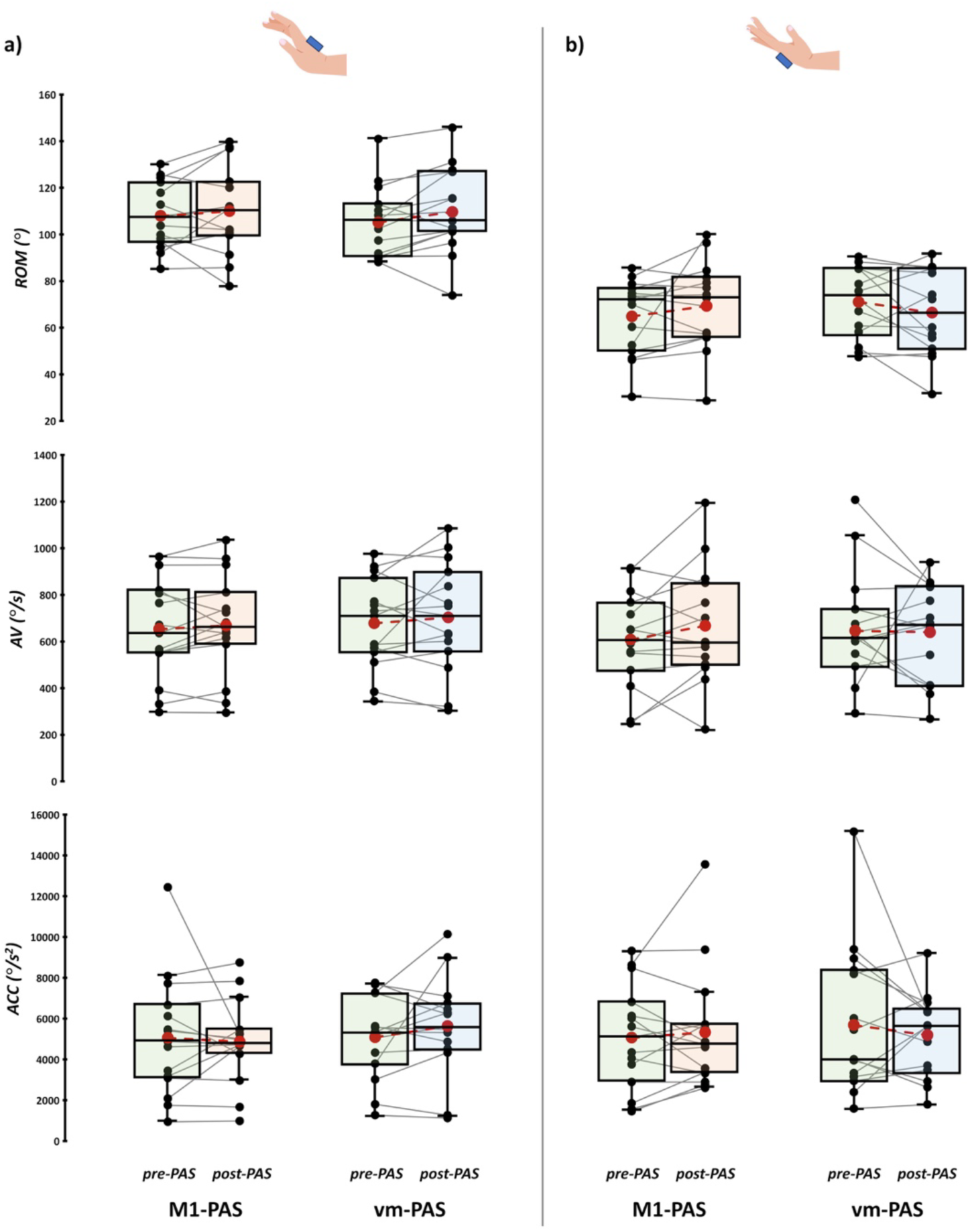
Paretic wrist’s voluntary movement assessment results. Results from voluntary movement assessment during paretic wrist extension (**a**) and flexion (**b**), before (green boxes) and after vm-PAS (blue boxes) and M1- PAS (orange boxes) administration. No significant effects were found for median ROM, AV, and ACC values. Red dots and lines indicate the means of the distributions. The center line denotes their median values. Black dots and grey lines show individual scores (N=15). The box contains the 25th to 75th percentiles of the dataset. Whiskers extend to the largest observation falling within the 1.5 * inter-quartile range from the first/third quartile.

### 3.3. Correlations between PAS modulation and patients’ clinical profile

We found a significant positive correlation between vm-PAS effects on CSE and stroke chronicity: namely, the more time elapsed from the stroke, the greater the enhancement of FDI MEPs, and thus the efficacy of the vm-PAS (*rho* = .53, *p* = .042). The same pattern was not found for ECR after M1-PAS (*rho* = -.06, *p* = .82). Similarly, also the PAS-unspecific CSE enhancement found for FCR did not correlate with stroke chronicity (vm-PAS: *rho* = -.37, *p* = .172; M1-PAS: *rho* = -.32, *p* = .247; **Figure 5**).

**Figure 5.**
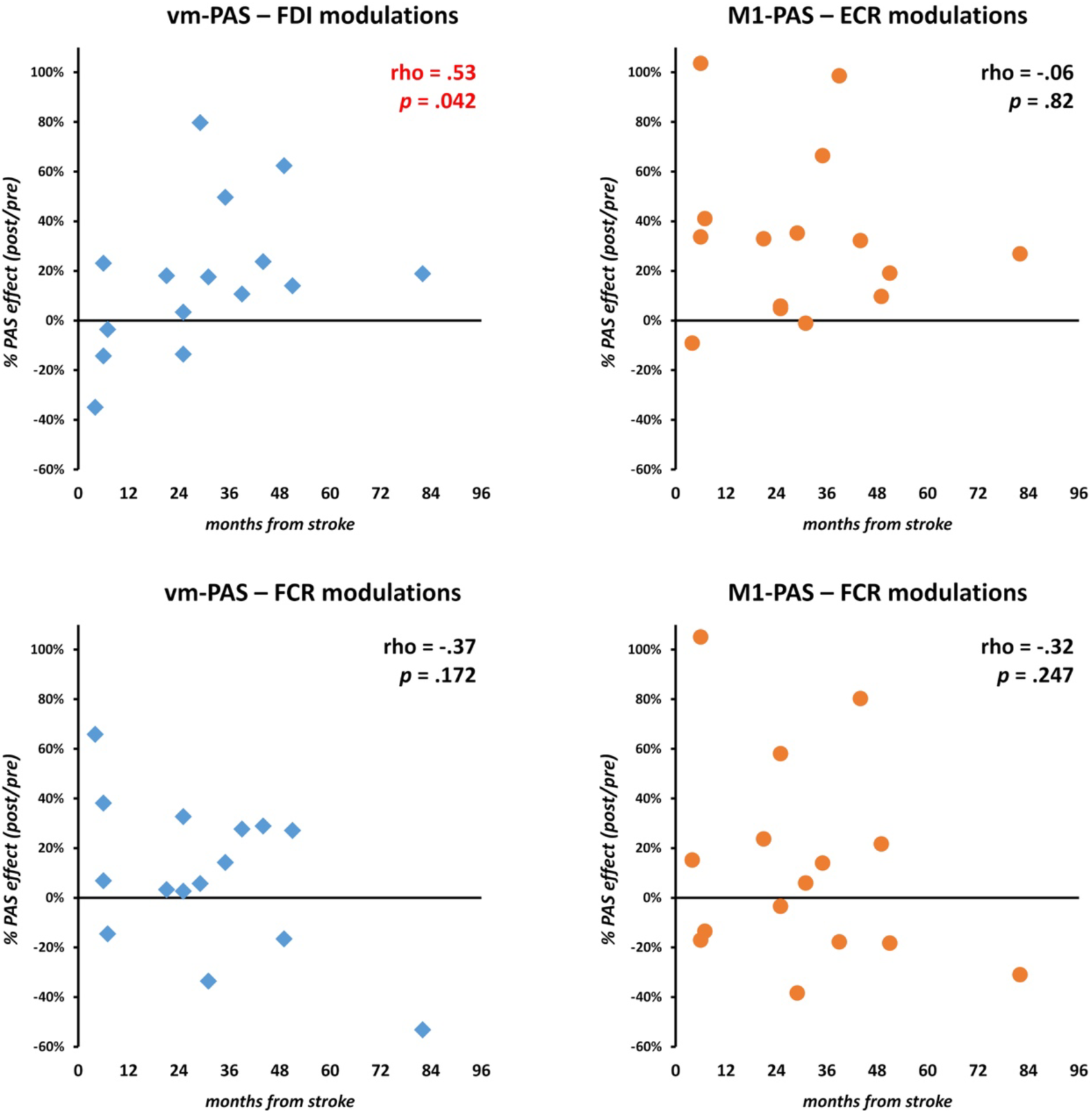
Correlation between CSE enhancement and stroke chronicity. Scatterplots between the percentage of MEP enhancement after PAS administration (left panels: vm-PAS, right panels: M1-PAS) and stroke chronicity (months elapsed from stroke) on the fifteen patients of our sample. Significant Spearman correlation was found only for vm-PAS administration.

Finally, FMA-UL and HGS scores, patients’ age and scholarity, M1 excitability, and voluntary wrist movements at baseline did not correlate with the corticospinal effects of both PAS protocols (all *rho*s < .44, all *p*s > .1; **Supplemental Figures 1-6**).

## 4. DISCUSSION

The present study shows the effectiveness of the vm-PAS in a cohort of chronic post-stroke patients with upper limb hemiparesis. This PAS protocol pairs TMS pulses over the ipsilesional M1 with visual stimuli of a reach-to-grasp action, enhancing, after its administration, CSE of the paretic limb in a muscle-specific fashion. At the same time, we also provide further support for the effectiveness of the standard M1-PAS that, however, modulates CSE with a different muscle-specific profile than vm-PAS.

The vm-PAS leverages adaptive mechanisms of visuomotor interaction mediated by the activation of the AON during the observation of the conditioned reach-to-grasp action (the so-called ‘motor resonance’ phenomenon- Craighero, 2024). Indeed, it is well known that during the observation of biological movements, M1 excitability is enhanced (Fadiga et al., 2005; Naish et al., 2014; Rizzolatti et al., 2014). This greater reactivity is thought to reflect the activation of mirror neuron populations located in the ventral premotor cortex, a key hub of the AON, which have direct connection with M1, in turn influencing motor system excitability during action observation (e.g., Cantarero et al., 2011; Catmur et al., 2011; Chiappini et al., 2024; de Beukelaar et al., 2016). Hence, the repeated observation of the hand action during the vm-PAS recruits this high-order sensorimotor network, likely in a somatotopic fashion (i.e., maximizing the vicarious recruitment of M1 population coding for the muscles involved in the observed grasping (Alaerts et al., 2009; Craighero, 2024; Fadiga et al., 2005; Rizzolatti, 2005)). Here, thanks to the time-locked coupling with the exogenous, TMS-driven activation of the ipsilesional M1, the vm-PAS promotes the induction of associative plasticity within the injured motor system, in turn enhancing CSE in the paretic limb after its administration. The vm-PAS likely strengthens cortico-cortical pathways of the AON – as suggested by recent evidence with cortico-cortical PAS on the healthy, (e.g., Chiappini et al., 2024; Turrini et al., 2024), but it does not require the integrity of the afferent sensory pathway from the affected limb for the induction of plasticity within M1, as indeed happens for the standard M1-PAS (Suppa et al., 2017). In fact, the vm-PAS protocol recruits the lesioned motor system through an indirect, visual-mediated pathway, usually spared in patients with hemiparesis. This evidence not only corroborates previous works in healthy individuals (Guidali et al., 2020, 2025; Guidali, Picardi, Gramegna, et al., 2023; Guidali & Bolognini, 2025), but it also documents the efficacy of the vm-PAS for enhancing motor cortex excitability in an injured motor network. Considering the effectiveness of the 25 ms ISI between vm-PAS paired stimulations, previous works on the healthy showed that it could reflect the recruitment of anticipatory mechanisms (Guidali & Bolognini, 2025; Maddaluno et al., 2020). Indeed, the action observation stimuli presented during the protocol are highly predictable, as patients repeatedly observed the same grasping stimulus for 15 minutes at a fixed frequency (i.e., every 5 sec). As the vm-PAS progresses, the AON can likely be engaged in advance, even before the actual view of the movement. This idea is supported by previous studies on the AON in both humans and primates, demonstrating action anticipation phenomena driven by the presentation of sufficient contextual cues about the movement to be observed (e.g., Aglioti et al., 2008; Qin et al., 2023; Urgesi et al., 2010).

An intriguing finding is the muscle-selective enhancement of ipsilesional M1 excitability, which further supports the contribution of the AON (e.g., Amoruso & Urgesi, 2016; Bunday et al., 2016; Cavallo et al., 2012; Gangitano et al., 2001; Guidali, Picardi, Franca, et al., 2023; Koch et al., 2010): the vm-PAS drives the mirror recruitment of the muscles involved in the observed grasping action, namely FDI and FCR, rather than the muscle primarily engaged in the reaching phase (ECR; (Anson et al., 2002; Montagna et al., 2005)) and plasticity in M1 is induced accordingly. It is well known that during reach-to-grasp movements, ECR does not act as an agonist but rather as a stabilizer, supporting the wrist throughout the action (Neumann, 2010). This suggests that during the observation of a grasping action, motor resonance might be more pronounced in FDI than in ECR, given that the former is one of the primary agonist muscles for this kind of movement. In turn, plasticity is induced in the motor system following the somatotopic features of the observed, PAS-conditioned action. Such muscle-specific modulation, could have been further maximized by attention. Indeed, during the vm-PAS session, patients were instructed to ‘*count the number of times the bottle was grasped by the hand*’ to the best of their ability, focusing their attention on the target of the visual movement, hence on the hand grasping the bottle, rather than the wrist movement. An additional factor favoring this sort of attentional capture is the ‘single-frame’ nature of the visual stimulus of the vm-PAS, depicting the reach-to-grasp movement that, unlike a video, does not show the entire action sequence but only the end of the action, that is the grasping phase. Considering the crucial role of the observer’s attention for motor resonance and AON functioning (e.g., Puglisi et al., 2017; Schuch et al., 2010), as well as for the PAS efficacy (Stefan et al., 2004), the focus of patients’ attention on the grasping phase of the observed action may have maximized the mirror activation of FDI and FCR representations in M1. This focus has, in turn, favored a muscle-specific enhancement of MEPs according to the final grasping phase of the observed action (Guidali, Picardi, Franca, et al., 2023).

Of interest are the complementary effects brought about by the M1-PAS. With this protocol, we replicate the MEP enhancement of ECR and FCR muscles found in previous studies using, as here, the exact version of the M1-PAS (Castel-Lacanal et al., 2007, 2009), but in a larger cohort of stroke patients [n=15 *vs.* 2 and 6 of Castel-Lacanal et al. (2007) and Castel-Lacanal et al. (2009), respectively]. At variance of the vm-PAS, the M1-PAS increases MEPs recorded from ECR, the muscle electrically stimulated during the PAS, and FCR without affecting MEPs from the FDI muscle. This result is at odds with the ‘topographical specificity’ of the electrical PAS: the excitability changes sustained by the PAS are limited to the muscles innervated by the peripheral afferents, which have been stimulated electrically (Stefan et al., 2000; Wolters et al., 2003). However, the PAS topographical specificity should be looked for more in the effect size of the induced effects rather than in the complete absence of PAS-induced facilitation in muscles innervated by nerves antagonist to that stimulated during the M1-PAS, as confirmed by several works (Carson & Kennedy, 2013; Suppa et al., 2017). In addition to the not-so-strict topographical specificity of the PAS, other considerations could justify the spread to the FCR muscle of the excitation induced by the PAS of the radial nerve and the cortical ECR hotspot. In several instances, FCR and ECR act synergistically. Even if FCR and ECR can act as antagonists to each other, such as in simple flexion-extension wrist movements, they have unique features that set them apart from other agonist-antagonist couples (Aguiar & Baker, 2018). For example, regarding their Ia innervation, which is likely involved in the M1-PAS, it has been questioned that the Ia-mediated reciprocal inhibition, a neurophysiological hallmark of the agonist-antagonist distinction, exists for the FCR and ECR in the human forearm (Pierrot-Deseilligny & Burke, 2012).

Summarizing: while the vm-PAS, using a visual afferent pathway to activate the AON, induces a more selective enhancement of CSE, which depends on an inner mirroring of the observed movement on which attention is put, the effects of the M1-PAS, using electrical nerve stimulation, seems to rely on the activation of synergic muscle representations. Notably, the responders *vs.* non-responders ratio at the two protocols aligns well with previous evidence showing PAS effectiveness in about the 60-80% of the tested sample (Guidali et al., 2020; Guidali & Bolognini, 2025; Minkova et al., 2019; Strube et al., 2015).

Of importance, PAS effects emerged after a single stimulation session in our sample of patients, who were in a chronic stage of illness (30±21 months from stroke, on average) and presented a persistent hand motor impairment. This evidence corroborates previous literature (Ferris et al., 2018; Palmer et al., 2018) and suggests PAS efficacy beyond the subacute stages of stroke recovery, where cortical plasticity is heightened (Baroni et al., 2024), rendering patients more responsive to rehabilitation interventions (Kwakkel et al., 2004). Noteworthy, only the neurophysiological improvements induced by the vm-PAS, but not those by the M1- PAS, positively correlate with the time elapsed from stroke: the more chronic the motor deficit, the more effective the vm-PAS is. Central adaptive mechanisms can modify the sensory information hierarchy in the face of peripheral sensory deficit, leading to the reliance on more robust sensory information for action. In the post-stroke chronic phase, the patient may thus develop an adaptation related to a visual reliance that engages the AON to compensate for the hand motor deficit. Namely, a somatosensory, proprioceptive deficit may cause a reorganization of sensory hierarchies, leading to greater dependence on visual input for motor execution. This increased reliance on vision during reaching-to-grasp movements likely amplifies the involvement of visuomotor circuits within the AON. Indeed, research suggests that in individuals with impaired proprioception or motor control, visual input compensates by reinforcing alternative pathways, including those in the AON, to guide movement execution (Bernard-Espina et al., 2021; Ma et al., 2022).

Concerning the SICI, we replicated the muscle-specific enhancement of CSE after both PAS protocols (see **Supplemental Analysis 1**), but without difference between conditioned and unconditioned MEPs, hence without modulating the SICI itself and indicating that they did not influence GABA-mediated inhibition (Butefisch, 2000; Hanajima et al., 1998), replicating previously evidence in patients (Castel-Lacanal et al., 2007; Ferris et al., 2018; Palmer et al., 2018; Quartarone et al., 2003).

Finally, the vm-PAS and the M1-PAS did not even have behavioral effects, at least concerning voluntary wrist movements and after a single protocol’s administration. This result is consistent with the need for multiple PAS sessions to observe clinically relevant behavioral improvements in stroke [(Baroni et al., 2024; Liang et al., 2024; Sui et al., 2021) but see Tarri et al. (2018) for controversial results]. An alternative, yet complementary, hypothesis for the null-results on voluntary wrist movements could be that patients’ lesion profiles may have restricted residual motor function of the hemiparetic limb, limiting the protocols’ capacity to translate neurophysiological changes into behavioral improvements.

One limitation of the present study is that it only includes patients with mild hemiparesis. This factor prevents the generalization of the result to patients with more severe motor impairment. Moreover, the sensory deficit was not evaluated in detail, which could have shed better light on the clinical profile of (M1-PAS) responders. Future studies should explore the applicability of PAS interventions in patients with varying degrees of motor impairment and consider the severity of sensory deficits for PAS outcomes and effectiveness. Furthermore, we did not have structural magnetic resonance imaging of all our patients’ brains, preventing us from running lesion mapping analysis. Likely, vm-PAS efficacy strongly depends on the sparing of AON parietal and frontal nodes (Molenberghs et al., 2012), and non-responders to vm-PAS may have lesions in these cortical areas. Our patients mainly present subcortical stroke (see **Table 1**), likely restricting the generalizability of our results to patients with a cortical stroke (Di Pino et al., 2014). Future studies should deepen this by mapping patients’ lesion profiles and correlating them with PAS effects. Finally, future studies could implement direct measurements of AON activation in patients, e.g., motor resonance or automatic imitation (e.g., Craighero et al., 2023; Guidali, Picardi, Gramegna, et al., 2023), to deepen vm-PAS functional substrates and the extent of its aftereffects. At the same time, future research could also explore the generalizability of vm-PAS results to other movements or more complex actions, using different movements to be conditioned during the protocol. In conclusion, our results provide the first evidence for the corticospinal efficacy of a visuomotor PAS protocol targeting the AON in post-stroke patients. Our findings suggest that the vm-PAS effectively induces short-term cross-modal (i.e., visuomotor) associative plasticity within the lesioned M1, paving the way for the development of tailored PAS based, e.g., on the action observation of specific movements and gestures, as well as the exploration of the possible synergic effects of this protocol with motor training or other conventional post-stroke rehabilitation therapies (Bolognini et al., 2016; Dimyan & Cohen, 2011).

## AUTHORSHIP CONTRIBUTION STATEMENT

MP, GG, and NB conceptualized and designed the experiment. MP and AC recruited the patients. MP and GG took part in data collection, analysis, and prepared the figures. GG, AC and NB supervised the experiment. MP, GG, and NB wrote the first draft of the manuscript. All authors discussed the results, revised the manuscript, and approved its final version.

## DATA AVAILABILITY STATEMENT

The data supporting the findings of this study will be publicly available on OSF (https://osf.io/xgn7f) upon publication.

## ACKNOWLEDGMENTS

We thank Francesca Crespi for her valuable help during data collection.

## SUPPLEMENTARY MATERIALS

Supplementary materials of the article contain: supplemental analysis on raw MEP amplitude during SICI assessment (**Supplemental Analysis 1**), patients’ scores in the Oxford Cognitive Scale (OCS) scores (**Supplemental Table 1**), skewness and kurtosis values of raw and log_10_-transformed MEPs recorded for the different muscles and assessments (**Supplemental Table 2**), LMM and rmANOVA results for the different analyses presented in the article (**Supplemental Tables 3-8**), and correlation plots between significant PAS-induced MEP modulation and (a) FMA-UL and HGS scores, (b) patients’ age and level of education, (c) individual rMT, (d) patients’ voluntary wrist movements assessment at baseline (**Supplemental Figures 1-6**).

