## Supplemental files for "Visuomotor paired associative stimulation enhances corticospinal excitability in post-stroke patients with upper-limb hemiparesis: A proof-of-principle study"

### – SUPPLEMENTARY MATERIALS –

This document contains the following Supplementary Materials:

- **Supplemental Analysis 1**
- **Supplemental Tables 1-8**
- **Supplemental Figures 1-6**

### SUPPLEMENTAL ANALYSIS 1

To assess whether CSE was still modulated during SICI assessment, we also conducted a linear mixed model (LMM; similar to the one run for CSE assessment at rest) on MEPs recorded during SICI. Visual inspection of the QQ-plot and skewness/kurtosis values showed that raw MEP amplitudes were not normally distributed SICI assessments. Base-10 log transformation made the distribution closer to normality (see **Supplemental Table 2**); hence, MEPs were transformed accordingly and then analyzed using LMM. Each muscle (ECR, FDI, and FCR) was analyzed separately with within-subjects factors 'PAS' (vm-PAS, M1-PAS), 'Time' (pre-PAS, post-PAS), 'MEP type' (conditioned, unconditioned), and their interaction as fixed effects; the intercept as the random one; and the patient as the cluster variable. For SICI, the within-subjects factor was included in the model.

Results showed significant effects for the main factor 'MEP type' for all our target muscles: unconditioned MEPs had a greater peak-to-peak amplitude than conditioned one (all *F*s > 50.62; all *p*s < .001; **SA – Tables 1-3**). Furthermore, we found a significant 'PAS' X 'Time' interaction only for ECR ($F_{1,1503} = 10.93$, $p < .001$) and FDI muscles ($F_{1,1503} = 10.15$, $p = .001$): i.e., regardless of being conditioned or unconditioned MEPs, their amplitude was enhanced after M1-PAS administration (all *t*s > 4.24, all *p*s < .001) and after vm-PAS (all *t*s > 4.69, all *p*s < .001), respectively (**SA Figure 1**). The triple interaction 'PAS' X 'Time' X 'MEP type' and the other double interactions did not reach statistical significance (all *F*s < 3.07; all *p*s > .08).

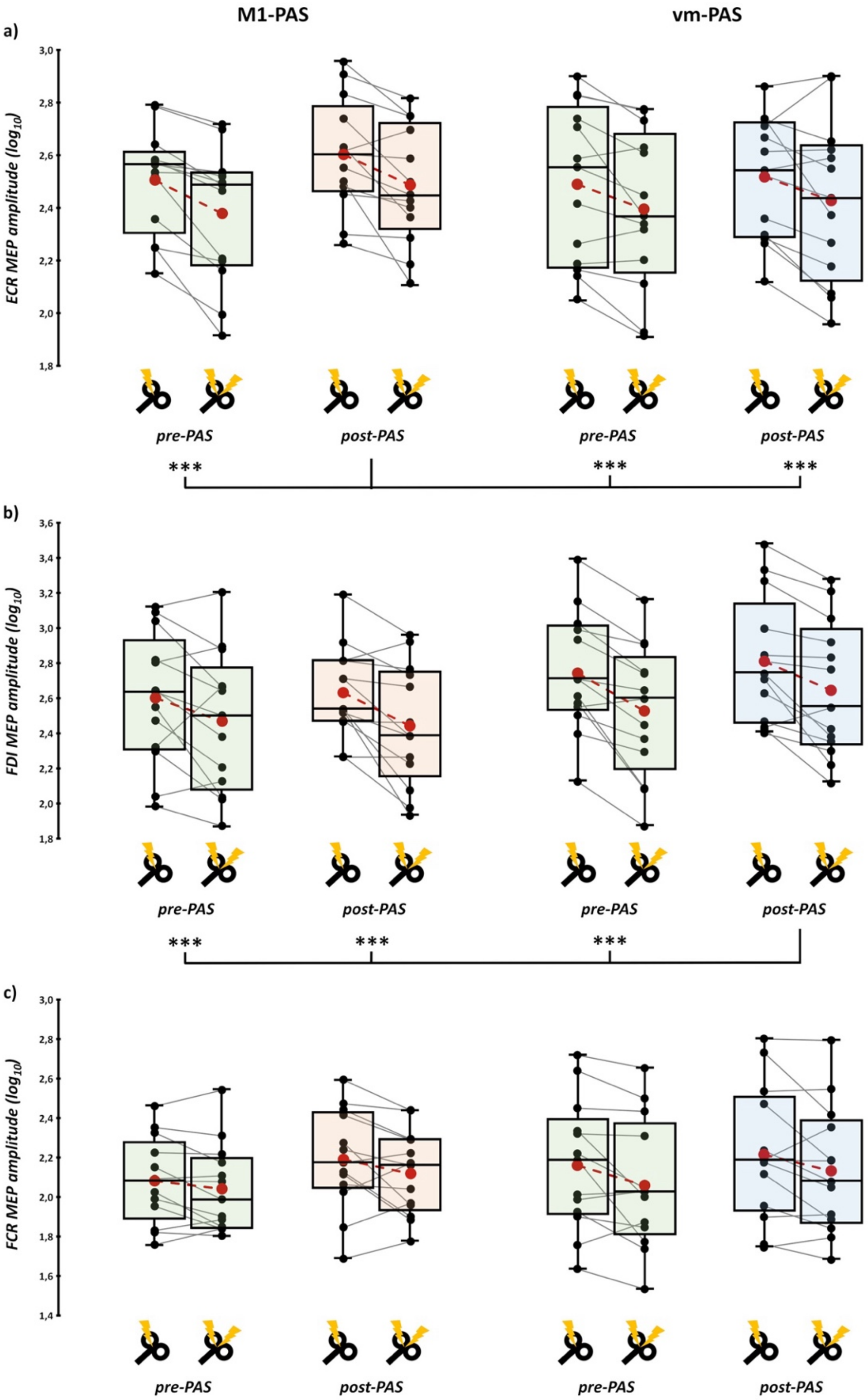


**SA - Figure 1.** *SICI results on MEPs.* LMM results from the SICI assessment for ECR (**a**), FDI (**b**), and FCR (**c**) muscles before (green boxes) and after vm-PAS (blue boxes) and M1-PAS (orange boxes) administration. Unconditioned MEPs had a greater peak-to-peak amplitude than conditioned one. Furthermore, in line with CSE patterns, their amplitude was enhanced after M1-PAS administration for ECR and after vm-PAS for FDI, regardless of being conditioned or unconditioned ones. Red dots and lines indicate the means of the distributions. The center line denotes their median values. Black dots and grey lines show individual values (N=13). The box contains the 25th to 75th percentiles of the dataset. Whiskers extend to the largest observation falling within the 1.5 * inter-quartile range from the first/third quartile. Asterisks indicate significant differences between conditions (* = p < .05, ** = p < .01; *** = p < .001; Bonferroni corrected).

| **ECR -** Fixed Effects Omnibus Tests | | | | | |
|---|---|---|---|---|---|
| *Factor* | *F* | | *df* | | *p* |
| **PAS** | 7.051 | | 1, 1503 | | .008 |
| **Time** | 34.053 | | 1, 1503 | | <.001 |
| **MEP type** | 87.820 | | 1, 1503 | | <.001 |
| **PAS X Time** | 10.932 | | 1, 1503 | | <.001 |
| **PAS X MEP type** | 2.051 | | 1, 1503 | | .152 |
| **Time X MEP type** | .135 | | 1, 1503 | | .713 |
| **PAS X Time X MEP type** | .221 | | 1, 1503 | | .638 |
| Parameter Estimates | | | | | |
| *Effect* | *Estimate* | *SE* | *df* | *t* | *p* |
| **Intercept** | 2.471 | .058 | 12 | 42.894 | <.001 |
| **vm-PAS - M1-PAS** | -.029 | .011 | 1503 | -2.655 | .008 |
| **pre - post** | -.065 | .011 | 1503 | -5.835 | <.001 |
| **unconditioned - conditioned** | .104 | .011 | 1503 | 9.371 | <.001 |
| **(vm-PAS - M1-PAS) X (pre - post)** | -.073 | .022 | 1503 | -3.306 | <.001 |
| **(vm-PAS - M1-PAS) X (unconditioned – conditioned)** | .032 | .022 | 1503 | 1.432 | .152 |
| **(pre - post) X (unconditioned – conditioned)** | .008 | .022 | 1503 | .368 | .713 |
| **(vm-PAS - M1-PAS) X (pre - post) X (unconditioned – conditioned)** | -.021 | .044 | 1503 | -.47 | .638 |

| **FDI -** Fixed Effects Omnibus Tests | | | | | |
|---|---|---|---|---|---|
| *Factor* | *F* | | *df* | | *p* |
| **PAS** | 113.055 | | 1, 1503 | | <.001 |
| **Time** | 11.629 | | 1, 1503 | | <.001 |
| **MEP type** | 153.769 | | 1, 1503 | | <.001 |
| **PAS X Time** | 10.15 | | 1, 1503 | | .001 |
| **PAS X MEP type** | .971 | | 1, 1503 | | .325 |
| **Time X MEP type** | .007 | | 1, 1503 | | .933 |
| **PAS X Time X MEP type** | 3.07 | | 1, 1503 | | .08 |
| Parameter Estimates | | | | | |
| *Effect* | *Estimate* | *SE* | *df* | *t* | *p* |
| **Intercept** | 2.620 | .085 | 12 | 30.724 | <.001 |
| **vm-PAS - M1-PAS** | .143 | .013 | 1503 | -2.655 | <.001 |
| **pre - post** | -.046 | .013 | 1503 | -5.835 | <.001 |
| **unconditioned - conditioned** | .167 | .013 | 1503 | 12.4 | <.001 |
| **(vm-PAS - M1-PAS) X (pre - post)** | .086 | .027 | 1503 | 3.186 | .001 |
| **(vm-PAS - M1-PAS) X (unconditioned – conditioned)** | -.027 | .027 | 1503 | -.985 | .325 |
| **(pre - post) X (unconditioned – conditioned)** | -.002 | .027 | 1503 | -.084 | .933 |

| | | | | | |
|---|---|---|---|---|---|
| **(vm-PAS - M1-PAS) X (pre - post) X (unconditioned – conditioned)** | .094 | .054 | 1503 | 1.752 | .08 |

| **FCR -** Fixed Effects Omnibus Tests | | | |
|---|---|---|---|
| ***Factor*** | ***F*** | ***df*** | ***p*** |
| **PAS** | 11.417 | 1, 1503 | <.001 |
| **Time** | 59.535 | 1, 1503 | <.001 |
| **MEP type** | 50.619 | 1, 1503 | <.001 |
| **PAS X Time** | 2.842 | 1, 1503 | .092 |
| **PAS X MEP type** | 1.684 | 1, 1503 | .195 |
| **Time X MEP type** | .117 | 1, 1503 | .733 |
| **PAS X Time X MEP type** | .817 | 1, 1503 | .366 |

| Parameter Estimates | | | | | |
|---|---|---|---|---|---|
| ***Effect*** | ***Estimate*** | ***SE*** | ***df*** | ***t*** | ***p*** |
| **Intercept** | 2.126 | .072 | 12 | 29.718 | <.001 |
| **vm-PAS - M1-PAS** | .036 | .011 | 1503 | 3.379 | <.001 |
| **pre - post** | -.083 | .011 | 1503 | -7.716 | <.001 |
| **unconditioned - conditioned** | .076 | .011 | 1503 | 7.115 | <.001 |
| **(vm-PAS - M1-PAS) X (pre - post)** | -.036 | .021 | 1503 | -1.686 | .092 |
| **(vm-PAS - M1-PAS) X (unconditioned – conditioned)** | -.028 | .021 | 1503 | -1.298 | .195 |
| **(pre - post) X (unconditioned – conditioned)** | -.007 | .021 | 1503 | -.342 | .733 |
| **(vm-PAS - M1-PAS) X (pre - post) X (unconditioned – conditioned)** | .039 | .043 | 1503 | .904 | .366 |

**SA – Tables 1-3.** Results from the LMMs conducted on $log_{10}$-transformed MEP amplitude during SICI.

| | 1 | 2 | 3 | 4 | 5 | 6a | 6b | 7a | 7b | 7c | 8 | 9a | 9b | 9c | 10a | 10b | 10c |
|---|---|---|---|---|---|---|---|---|---|---|---|---|---|---|---|---|---|
| **S01** | 4 | 3 | 4 | 4 | 15 | 3 | 4 | 50 | 0 | 0 | 9* | 4 | 3* | 4 | 6 | 6 | 13 |
| **S02** | 4 | 3 | 4 | 4 | 10* | 3 | 4 | 41* | 1* (r) | * | 12 | 2* | 2* | 4 | 6 | 6 | 13 |
| **S03** | 4 | 3 | 4 | 3* | 15 | 3 | 4 | 47 | 0 | 0 | 12 | 0* | 0* | 4 | 6 | 6 | 13 |
| **S04** | 4 | 3 | 4 | 4 | 11* | 3 | 4 | 47* | 0 | 0 | 12 | 0* | 3* | 4 | 6 | 6 | 13 |
| **S05** | 4 | 3 | 4 | 4 | 14 | 3 | 4 | 46* | 0 | 0 | 12 | 2* | 4 | 4 | 6 | 6 | 13 |
| **S06** | 4 | 3 | 4 | 4 | 14 | 3 | 4 | 48 | 0 | 0 | 12 | 3* | 3* | 3* | 6 | 6 | 13 |
| **S07** | 0* | 2* | 4 | 4 | 2* | 3 | 3* | 45* | 0 | * | 9* | 0* | 4 | 4 | 6 | 6 | 13 |
| **S08** | 4 | 3 | 4 | 4 | 15 | 3 | 4 | 49 | 0 | 0 | 12 | 4 | 4 | 4 | 6 | 6 | 11* |
| **S09** | 4 | 3 | 4 | 4 | 15 | 3 | 4 | 42* | 0 | * | 12 | 3* | 3* | 4 | 5* | 6 | 10* |
| **S10** | 3* | 3 | 4 | 4 | 15 | 3 | 4 | 47* | 0 | * | 9* | 0* | 0* | 2* | 6 | 6 | 12* |
| **S11** | 3* | 3 | 4 | 4 | 3* | 0* | 4 | 47 | 0 | 0 | 10* | 0* | 0* | 4 | 6 | 6 | 5* |
| **S12** | 1* | 3 | 2* | 3* | 3* | 2* | 0* | 47 | 1* (r) | 0 | 0* | 0* | 0* | 0* | 6 | 6 | 13 |
| **S13** | 4 | 3 | 4 | 4 | 15 | 3 | 4 | 50 | 0 | 0 | 12 | 0* | 3* | 4 | 6 | 6 | 13 |
| **S14** | 4 | 3 | 4 | 4 | 15 | 3 | 4 | 48 | 1* (r) | 0 | 12 | 4 | 4 | 4 | 6 | 6 | 13 |
| **S15** | 4 | 3 | 4 | 4 | 15 | 3 | 4 | 49 | 0 | 0 | 12 | 1* | 3* | 4 | 6 | 6 | 11* |

**Supplemental Table 1.** Patients' Oxford Cognitive Scale (OCS) scores in the different items. OCS items are reported in columns. 1. pictures naming (max:4), 2. semantics (max:3), 3. orientation (max:4), 4. visual field (max:4), 5. reading (max:15), 6a. numbers writing (max:3), 6b-calculations (max:4), 7a-broken hearts (max:50), 7b-egocentric neglect, 7c-allocentric neglect, 8-praxis (max:12), 9a-recall & recognition (max:4), 9b-episodic memory (max:4), 9c-delay recall recognition (max:4), 10a-executive task – circles (max:6), 10b-executive task – triangles (max:6), 10c-executive task – mixed (max:13). We indicated with * the pathological items, adjusting for age, educational level and sex.

| Task | Muscle | raw MEPs | | $log_{10}$ MEPs | |
|---|---|---|---|---|---|
| | | skewness | kurtosis | skewness | kurtosis |
| **corticospinal excitability at rest** | ***ECR*** | 1.714 | 7.43 | -.358 | -.516 |
| | ***FDI*** | 1.138 | .765 | -.131 | -.775 |
| | ***FCR*** | 1.504 | 2.295 | .172 | -1.104 |
| **SICI** | ***ECR*** | 1.363 | 2.995 | -.301 | -.437 |
| | ***FDI*** | 1.808 | 3.377 | -.041 | -.601 |
| | ***FCR*** | 2.739 | 9.76 | .457 | -.176 |

**Supplemental Table 2.** Skewness and kurtosis values of raw and $log_{10}$-transformed MEPs recorded for the different muscles and assessments.

### ECR

**Fixed Effects Omnibus Tests**

| *Factor* | *F* | *df* | *p* |
|---|---|---|---|
| **PAS** | .728 | 1, 1734 | .394 |
| **Time** | 21.854 | 1, 1734 | <.001 |
| **PAS X Time** | 47.715 | 1, 1734 | <.001 |

**Parameter Estimates**

| *Effect* | *Estimate* | *SE* | *df* | *t* | *p* |
|---|---|---|---|---|---|
| **Intercept** | 2.589 | .069 | 14 | 37.562 | <.001 |
| **vm-PAS - M1-PAS** | .008 | .009 | 1734 | .853 | .394 |
| **pre - post** | .041 | .009 | 1734 | 4.675 | <.001 |
| **(vm-PAS - M1-PAS) X (pre - post)** | -.122 | .018 | 1734 | -6.908 | <.001 |

### FDI

**Fixed Effects Omnibus Tests**

| *Factor* | *F* | *df* | *p* |
|---|---|---|---|
| **PAS** | 14.863 | 1, 1734 | <.001 |
| **Time** | 6.573 | 1, 1734 | .01 |
| **PAS X Time** | 15.769 | 1, 1734 | <.001 |

**Parameter Estimates**

| *Effect* | *Estimate* | *SE* | *df* | *t* | *p* |
|---|---|---|---|---|---|
| **Intercept** | 2.836 | .089 | 14 | 31.824 | <.001 |
| **vm-PAS - M1-PAS** | .033 | .009 | 1734 | 3.855 | <.001 |
| **pre - post** | .022 | .009 | 1734 | 2.564 | .01 |
| **(vm-PAS - M1-PAS) X (pre - post)** | .068 | .017 | 1734 | 3.971 | <.001 |

**FCR**

| Fixed Effects Omnibus Tests | | | | | |
|---|---|---|---|---|---|
| ***Factor*** | ***F*** | | ***df*** | ***p*** | |
| **PAS** | 55.02 | | 1, 1734 | <.001 | |
| **Time** | 5.72 | | 1, 1734 | .017 | |
| **PAS X Time** | .161 | | 1, 1734 | .688 | |
| **Parameter Estimates** | | | | | |
| ***Effect*** | ***Estimate*** | ***SE*** | ***df*** | ***t*** | ***p*** |
| **Intercept** | 2.357 | .084 | 14 | 27.986 | <.001 |
| **vm-PAS - M1-PAS** | .069 | .009 | 1734 | 7.418 | <.001 |
| **pre - post** | .022 | .009 | 1734 | 2.392 | .017 |
| **(vm-PAS - M1-PAS) X (pre - post)** | -.007 | .019 | 1734 | -.401 | .688 |

**Supplemental Tables 3-5.** Results from the LMMs conducted on $log_{10}$-transformed MEP amplitude during corticospinal excitability assessment.

| *Muscle* | *Factor* | *F* | *df* | *p* | $\eta_p^2$ |
|---|---|---|---|---|---|
| | **PAS** | .01 | 1, 12 | .92 | .001 |
| **ECR** | **Time** | .064 | 1, 12 | .805 | .005 |
| | **PAS X Time** | .745 | 1, 12 | .405 | .058 |
| | **PAS** | .114 | 1, 12 | .742 | .009 |
| **FDI** | **Time** | .195 | 1, 12 | .667 | .016 |
| | **PAS X Time** | .161 | 1, 12 | .695 | .013 |
| | **PAS** | .516 | 1, 12 | .486 | .041 |
| **ECR** | **Time** | .67 | 1, 12 | .429 | .053 |
| | **PAS X Time** | .021 | 1, 12 | .886 | .002 |

**Supplemental Tables 6.** Results from the rmANOVAs conducted for SICI assessment.

### EXTENSION MOVEMENTS

| *Variable* | *Factor* | *F* | *df* | *p* | $\eta_p^2$ |
|---|---|---|---|---|---|
| **AV** | **PAS** | 2.964 | 1, 14 | .107 | .175 |
| | **Time** | 1.24 | 1, 14 | .284 | .081 |
| | **PAS X Time** | .054 | 1, 14 | .82 | .004 |
| **ROM** | **PAS** | .913 | 1, 14 | .356 | .061 |
| | **Time** | 3.511 | 1, 14 | .082 | .201 |
| | **PAS X Time** | .454 | 1, 14 | .511 | .031 |
| **ACC** | **PAS** | .769 | 1, 14 | .395 | .052 |
| | **Time** | .511 | 1, 14 | .487 | .035 |
| | **PAS X Time** | .689 | 1, 14 | .421 | .047 |

### FLEXION MOVEMENTS

| *Variable* | *Factor* | *F* | *df* | *p* | $\eta_p^2$ |
|---|---|---|---|---|---|
| **AV** | **PAS** | .192 | 1, 14 | .668 | .015 |
| | **Time** | .167 | 1, 14 | .69 | .013 |
| | **PAS X Time** | 1.321 | 1, 14 | .271 | .092 |
| **ROM** | **PAS** | .119 | 1, 14 | .735 | .009 |
| | **Time** | .008 | 1, 14 | .931 | .001 |
| | **PAS X Time** | 2.476 | 1, 14 | .14 | .16 |
| **ACC** | **PAS** | .615 | 1, 14 | .447 | .045 |
| | **Time** | .208 | 1, 14 | .656 | .016 |
| | **PAS X Time** | 1.71 | 1, 14 | .214 | .116 |

**Supplemental Tables 7-8.** Results from the rmANOVAs conducted for voluntary wrist movements assessment.

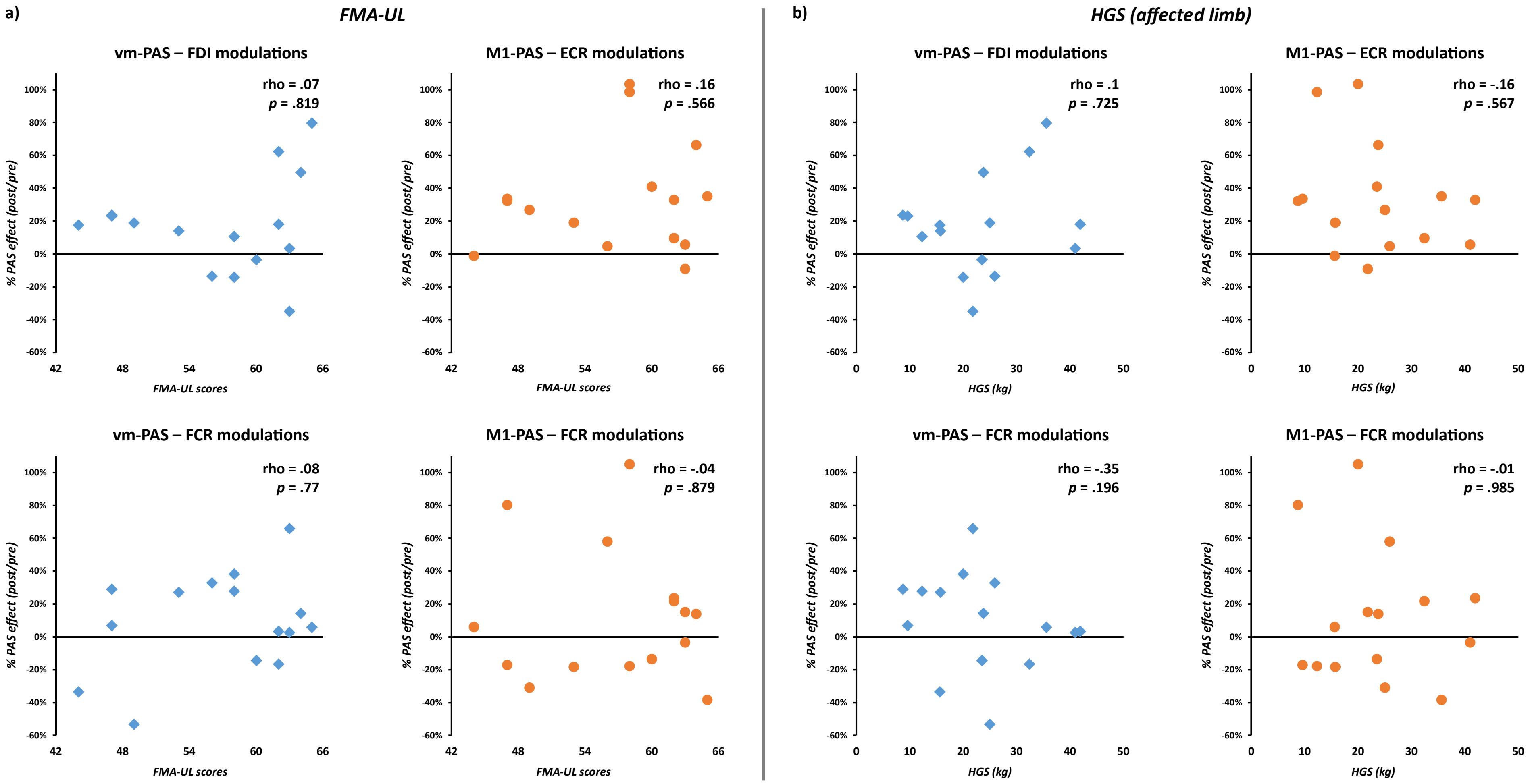


**Supplemental Figure 1.** Correlations between significant PAS-induced MEP modulation and FMA-UL (**a**) and HGS (**b**) scores.

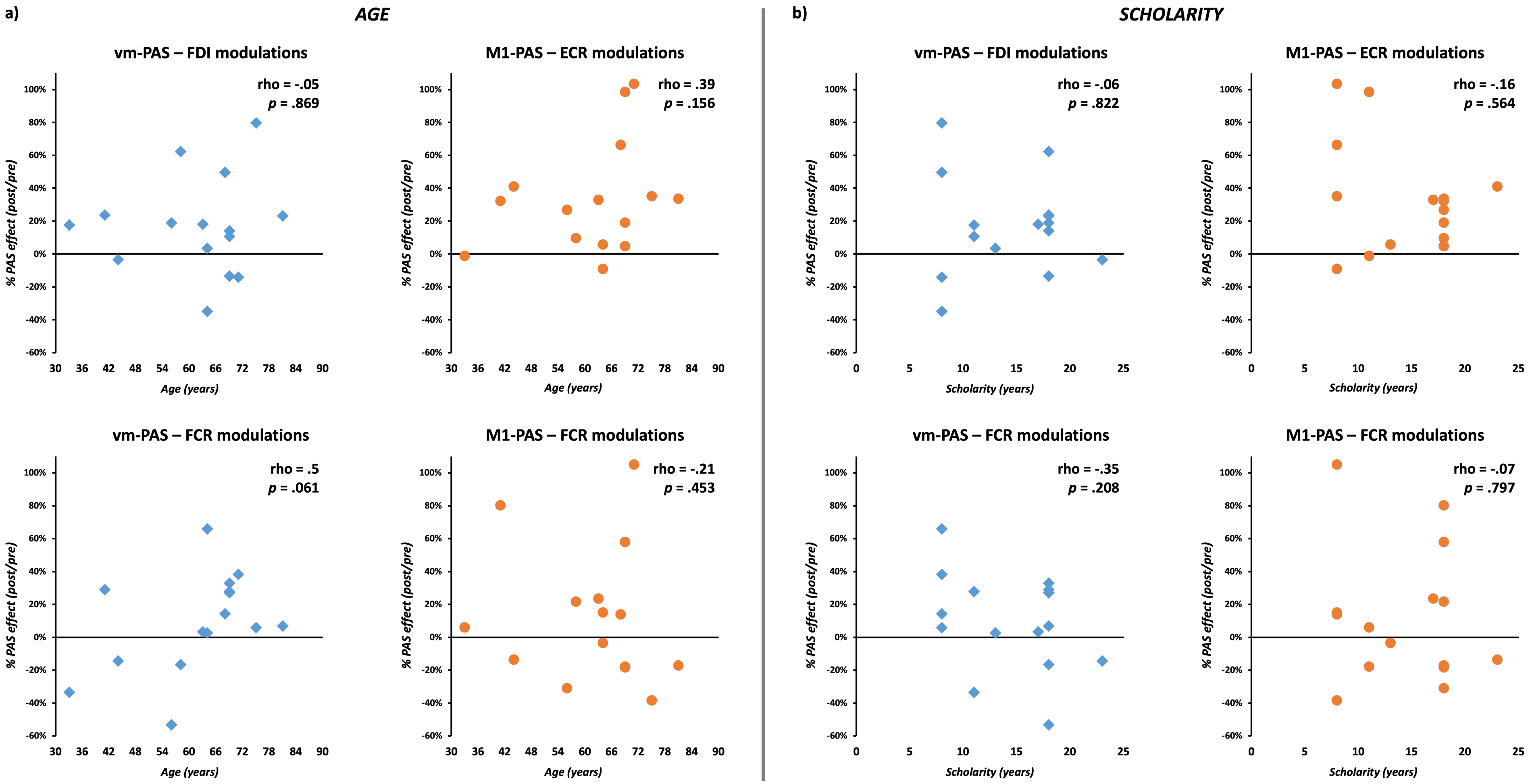


**Supplemental Figure 2.** Correlations between significant PAS-induced MEP modulation and patients' age (**a**) and scholarity (**b**).

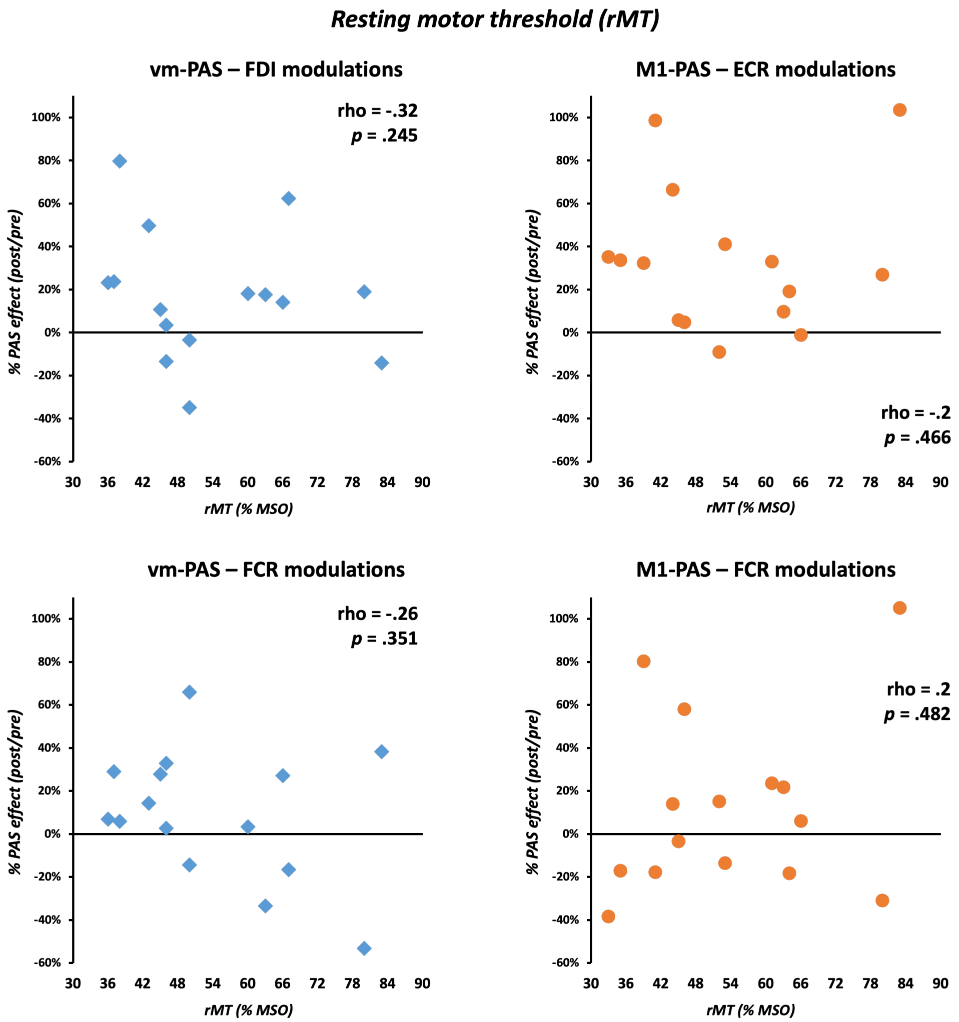


**Supplemental Figure 3.** Correlations between significant PAS-induced MEP modulation and patients' resting motor threshold (rMT).

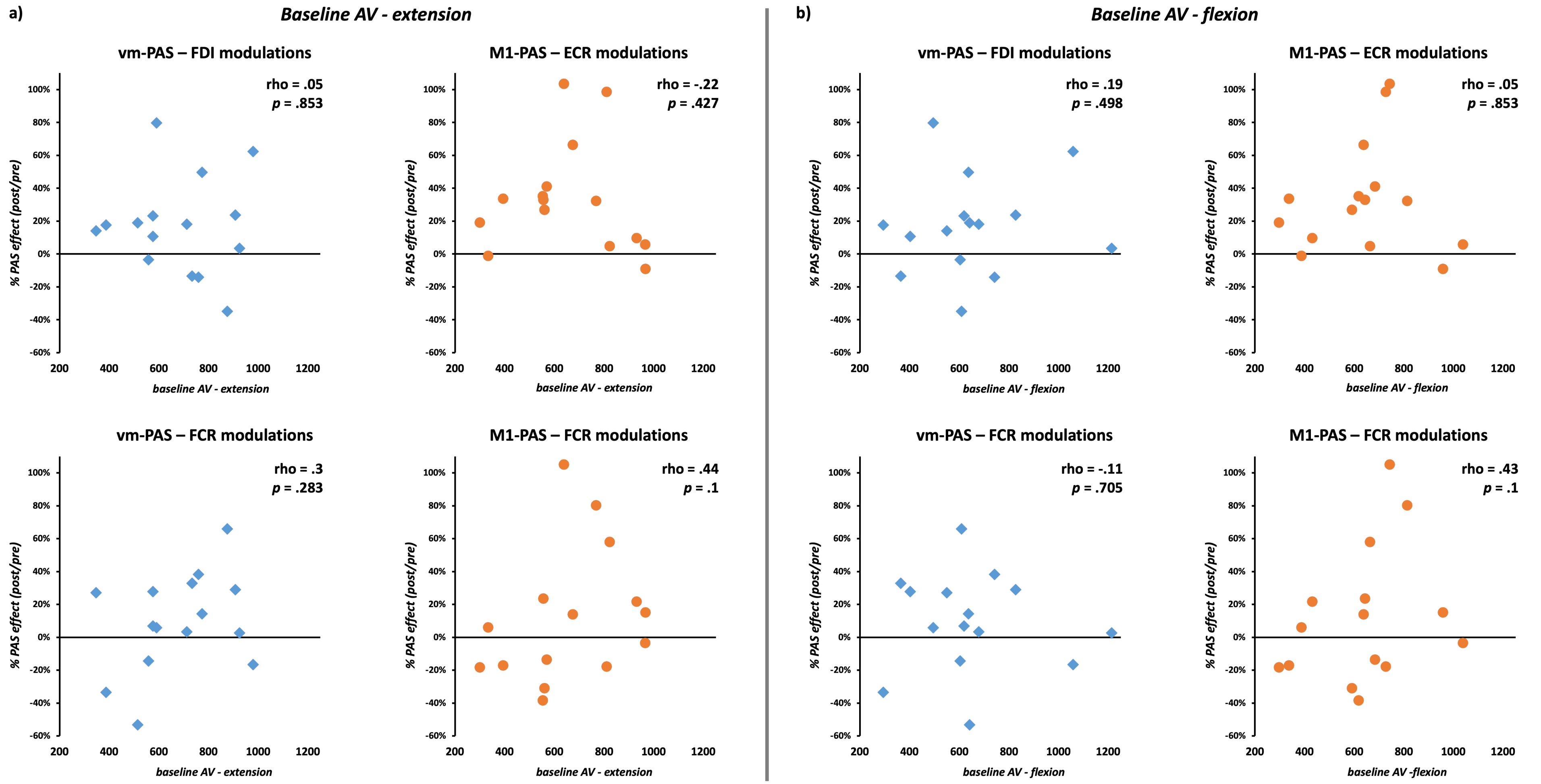


**Supplemental Figure 4.** Correlations between significant PAS-induced MEP modulation and patients' voluntary wrist movements (extension – **a**; flexion, **b**) angular velocity (AV) values during baseline assessments.

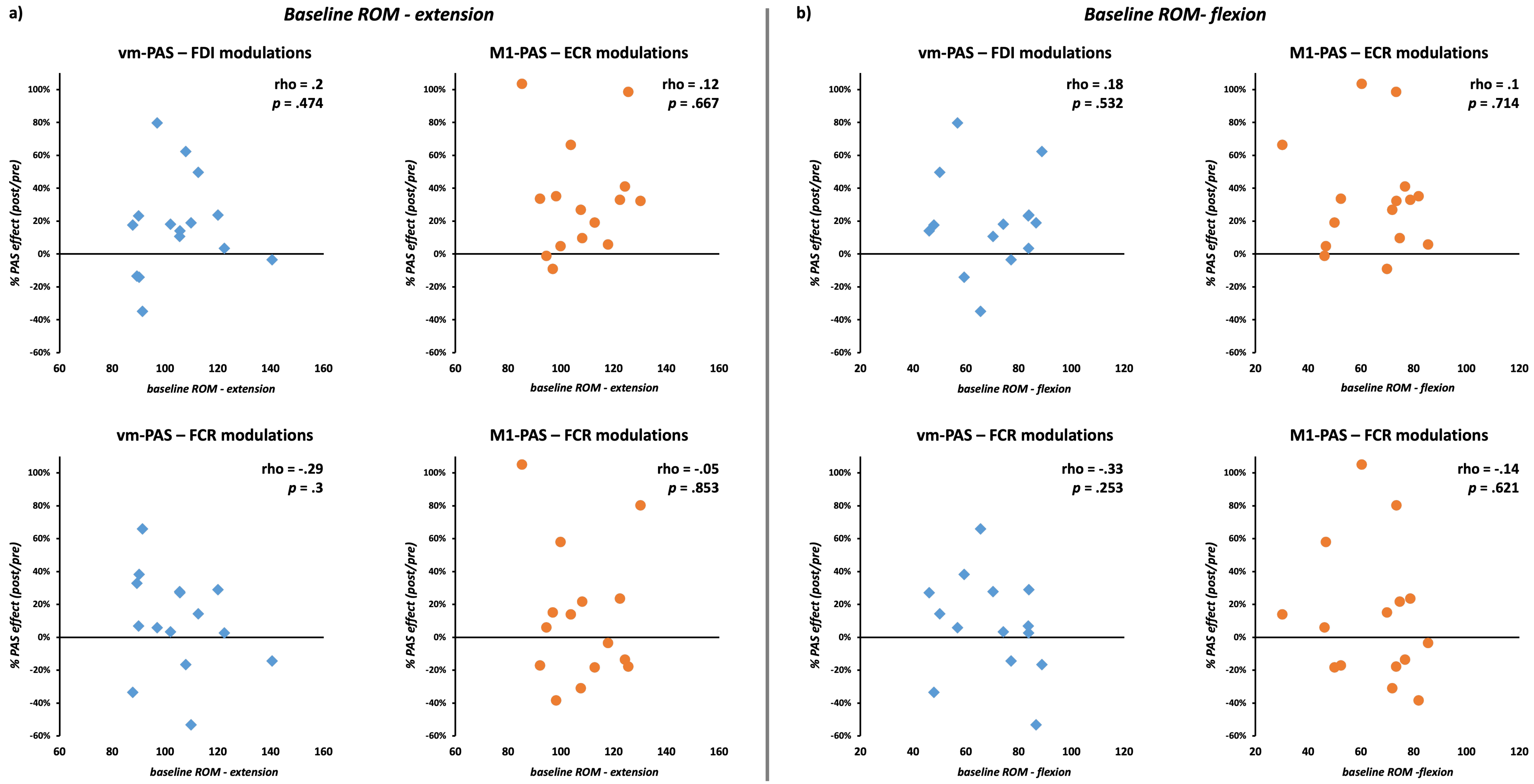


**Supplemental Figure 5.** Correlations between significant PAS-induced MEP modulation and patients' voluntary wrist movements (extension – **a**; flexion, **b**) range of motion (ROM) during baseline assessments.

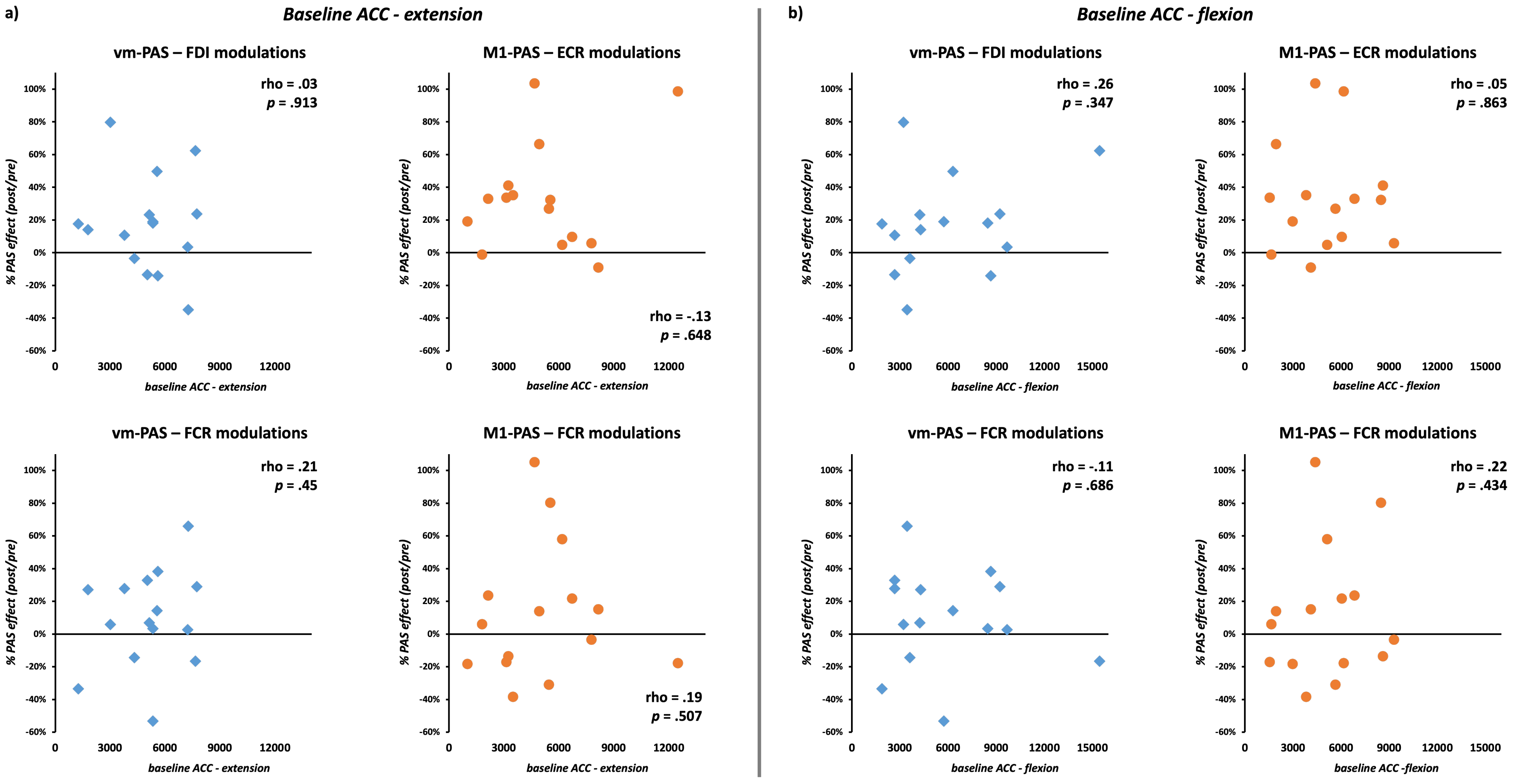


**Supplemental Figure 6.** Correlations between significant PAS-induced MEP modulation and patients' voluntary wrist movements (extension – **a**; flexion, **b**) acceleration (ACC) during baseline assessments.
